# Detecting intervention-specific change in soft tissue mobility aligned with regional pain through optically measured skin surface strains

**DOI:** 10.1101/2025.07.31.25332529

**Authors:** Anika R. Kao, M. Terry Loghmani, Gregory J. Gerling

## Abstract

**Purpose:** Soft tissue manipulation (STM) is a widely used massage-based intervention for myofascial pain, yet its effects on soft tissue mobility are primarily assessed through subjective clinical observations. Mechanical changes underlying symptom relief therefore remain unclear. No objective gold standard exists; current methods either measure stiffness in small regions, missing lateral mobility between fascial layers, or rely on clinical palpation. Optical measurement of tissue mobility from the skin surface offers an objective approach compatible with clinical practice.

**Methods:** Digital image correlation captured skin surface deformation during hands-on assessment of the cervicothoracic region. Nineteen participants underwent a standardized STM protocol. Tissue mobility was measured immediately before and after intervention, considering tissue pull direction (superior vs. inferior) and bilateral anatomy (left vs. right). Eleven strain-based biomarkers evaluated tissue glide and deformation.

**Results:** Several biomarkers changed significantly following intervention, with maximum pull increasing 3.4 mm. STM produced tissue mobility changes in nearly all treated participants, with 88% improving on at least one body side. Among participants with baseline bilateral pain asymmetries, 90% showed greater mobility gains on their more painful side. Bilateral mobility asymmetries were observed in 53% of participants, eight of whom reported pain asymmetries; in all cases, the less mobile side corresponded to the more painful side.

**Conclusion:** Optically derived tissue glide and deformation measures can objectively detect soft tissue mobility changes immediately following STM. Mobility gains align with self-reported pain, supporting the clinical relevance of strain-based biomarkers and their potential to complement clinical assessment of myofascial restriction and therapeutic response.

## Introduction

Myofascial pain, a prevalent and debilitating condition, is characterized by localized discomfort, tenderness, and restricted mobility of the myofascial tissue, often associated with the presence of myofascial trigger points (MTrPs) [1, 2]. These hyperirritable nodules within taut bands of skeletal muscle can lead to pain referral, muscle stiffness, and impaired function, significantly affecting quality of life [3–5]. To alleviate pain and restore soft tissue mobility, clinicians commonly employ soft tissue manipulation (STM) interventions, including techniques such as therapeutic massage, myofascial release, and targeted trigger point therapy [6–9]. These interventions are hypothesized to enhance fascial and muscle gliding, reduce local tissue restrictions, and modulate nociceptive input, thereby decreasing associated pain [10, 11]. Despite their widespread use, the immediate effects of STM are typically evaluated using subjective measures, such as patient-reported pain intensity or perceived improvement, which are often nonspecific and qualitative in nature [12, 13]. While reductions in self-reported pain and headache intensity have been documented following STM [14–16], such measures provide limited insight into the underlying biomechanical changes responsible for symptom relief. Consequently, there is a critical need for objective, quantitative methods capable of assessing soft tissue properties with high precision to elucidate the mechanical effects of hands-on intervention and the mechanisms by which they induce functional improvements [17].

The gold standard for assessing soft tissue mobility is clinician judgement informed by manual and instrument-assisted palpation to judge tenderness, hardness, restrictions, inflammation, and multidirectional glide [18, 19]. As such, standardization in assessment remains difficult and dependent on clinician experience and participant self-report [20]. On the other hand, tools to make objective measurements such as myotonometry and shear wave elastography (SWE) attempt to characterize tissue stiffness [21–24], but often sample only small regions and can be sensitive to probe alignment and tissue anisotropy [25]. Moreover, stiffness-based measures, such as those derived from algometer and myotonometer devices, do not capture the lateral mobility of soft tissues. Lateral mobility reflects the ability of fascial layers to glide relative to one another, a process that may be facilitated by hyaluronan within the extracellular matrix [26–28]. In animal models, both controlled injury and movement restrictions have been shown to limit lateral mobility, highlighting the critical role of inter-layer glide in maintaining normal tissue mechanics [29]. Investigating these dynamics *in vivo* in awake humans is essential, as cadaveric models cannot account for active muscle co-activation [30]. Although emerging ultrasound approaches (*e.g.,* anisotropy, shear-wave speed, Doppler flow) show promise for assessing tissue mechanics [31], reliable, region-wide measures of lateral mobility during clinically relevant manipulations remain lacking.

Assessing tissue glide and deformation from the skin surface provides additional insight into how bulk tissue responds to applied forces, as skin and tissues move differently depending on how pressure or displacement is applied. Capturing this motion can further understanding of the functional restrictions that contribute to pain. Touch-contact interactions have been explored across a variety of fields of study, using approaches that differ in resolution, scale, and complexity. For example, 2D and 3D positions of body segments can be tracked without markers using image analysis tools such as DeepLabCut, or through electromagnetic (Polhemus), stereo-infrared (Leap Motion), and structured-light (Microsoft Kinect) systems [32–39]. These marker-less approaches allow measurement of limb and hand movements without attaching sensors to the body, in contrast to systems like Vicon, which require physical markers on anatomical landmarks [40–42]. While these methods can effectively quantify movements of hands and limbs by tracking skeletal positions, a distinct task is the capture of lateral stretch and motion within the skin. In this regard, prior efforts have imaged skin surface movement, often at the fingertip against rigid glass or elastic surfaces, to study slip and localized deformation [43–47]. Such methods typically rely on disparity mapping to obtain 3D positions at discrete time points, but they cannot reveal how tissues stretch continuously over time, also known as strain. To measure continuous tissue stretch and strain over larger regions, techniques such as 3D digital image correlation (3D-DIC) or optical coherence tomography (OCT) are more suitable. OCT is generally limited to small imaging planes, spanning only a few millimeters [48, 49], whereas 3D-DIC with multiple cameras can combine multiple surfaces to avoid occlusions, cover tens of centimeters, and evaluate lateral skin stretch across broader anatomical regions [48–52]. Using this approach, we have developed strain-based biomarkers derived from skin surface deformation measurements. In human participants, these biomarkers were able to detect bilateral asymmetries in tissue mobility within an individual, which aligned with self-reported pain in 84.2% of cases [53].

Expanding on this prior work, the present study evaluates whether the strain-based biomarkers distinguish changes in tissue mobility before and after an established soft tissue manipulation intervention protocol. To maximize the likelihood of capturing measurable effects, the intervention was delivered by an expert clinician using both manual and instrument-assisted techniques. We hypothesized, first, that the biomarkers would be sufficiently sensitive to detect changes in tissue mobility immediately before and after the STM intervention, providing an objective complement to subjective reports of clinical improvement. Second, we expected that participants with bilaterally asymmetric pain would exhibit larger changes in tissue mobility on their more painful body side, consistent with greater initial restriction and treatment response. While no objective gold standard exists for assessing soft tissue mobility, optical measurement of soft tissue mobility from the skin surface offers a quantitative approach to complement clinical assessments of myofascial restriction and therapeutic response.

## Materials and Methods

This study optically captures the skin’s deformation in response to a clinician’s lateral stretch used to assess tissue of the cervicothoracic region. Skin surface mobility is quantified immediately before and after an STM intervention using 3D digital image correlation, synchronized input from three camera angles, and biosafe skin speckling techniques. From raw DIC data, eleven strain-based biomarkers are derived to evaluate tissue glide (how far tissue moves in response to loading) and tissue deformation (how much tissue stretches or compresses under that load). For the nineteen recruited participants, we examine changes given factors of intervention (pre vs. post), and asymmetries given bilateral anatomy (left vs. right body sides) and skin pull direction (superior vs. inferior relative to the participant’s back).

### Soft Tissue Manipulation Stretch Assessment

In the standardized assessment protocol, the clinician applied a controlled pull to the tissue at a 45-degree angle relative to the surface of the myofascial plane. This maneuver is guided by the clinician’s own tactile feedback toward a consistent target force rather than a fixed pull distance or speed. Static pressure was applied in a linear direction, horizontally along the myofascial plane, at a steady ramp-up rate of ∼3 seconds to maximum pressure. The maneuver was applied bilaterally (left and right body sides) and in multiple directions (superior towards the neck, and inferior away from the neck) from a central reference point, such as a tender spot [21, 54] (Fig. 1(a)). Clinically relevant factors in this assessment include the spatial distance and velocity of force propagation from the point of application, the point of force magnitude and deformation at which discomfort (if any) is reached, the magnitude of force required to reach maximum tissue stretch, and any identified restrictions or barriers to fascial motion (*e.g.,* adhesions, edema, scar). Tissue impaired with MTrPs restricts glide [3, 4], which may noticeably shorten the distance over which an applied pull propagates (Fig. 1(b, c)). Following STM intervention, one might expect to observe an increase in tissue mobility as associated with an increase in the glide of superficial skin and subcutaneous tissues over deeper muscle structures [55] (Fig. 1(c, d)).

**Fig. 1.**
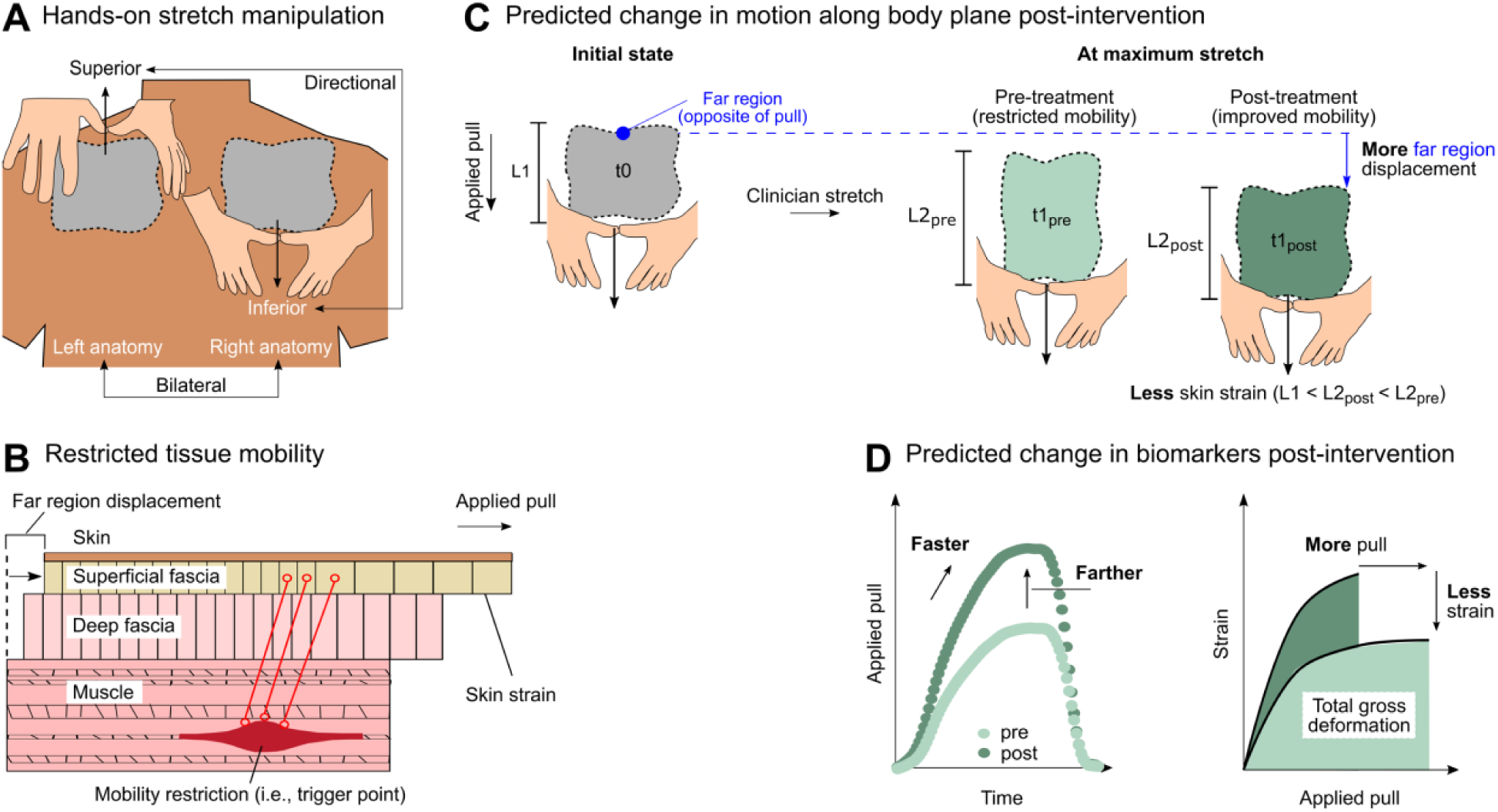
Conceptual overview of STM stretch assessment and predicted changes to soft tissue mobility following an STM intervention. (a) Schematic representation of STM stretch assessment, applied bilaterally (left and right sides of the body) and directionally (superior and inferior pull directions) in the cervicothoracic region. (b) Cross-sectional schematic of tissue layers (skin, superficial fascia, deep fascia, and muscle) highlighting restricted tissue mobility due to a myofascial trigger point, which resists a clinician-applied pull, thereby reducing tissue glide and propagation distance. (c) Predicted changes in motion along the body plane pre- and post-intervention. Pre-intervention (light green), restricted tissue is characterized by limited glide and increased deformation under clinician-applied pull. Post-intervention (dark green), tissue is anticipated to exhibit improved mobility with increased glide and reduced deformation. (d) Predicted changes in biomarker behavior pre- and post-intervention. Post-intervention (dark green), tissues are expected to demonstrate reduced resistance, allowing clinicians to pull faster and farther, with decreased reliance on tissue deformation (strain)

Force was not directly instrumented during this maneuver, as doing so would require a physical interface between the clinician’s hand and the participant’s skin, altering the tactile feedback central to this assessment [56].

### Soft Tissue Manipulation Intervention

To better control consistency and maximize the likelihood of measurable effects, a single clinician delivered the STM intervention to all applicable study participants. The total intervention time was 15 minutes, which included techniques both manual (5 minutes) and instrument-assisted (IASTM; 8 minutes) (Fig. 2(b)), followed by a 2 minute wrap-up to gradually taper off applied force. All aspects of the STM intervention remained within each participant’s level of comfort. Emollient was applied regionally to each participant’s skin to facilitate smooth and effective gliding of clinician contact and was removed afterward with alcohol wipes.

**Fig. 2.**
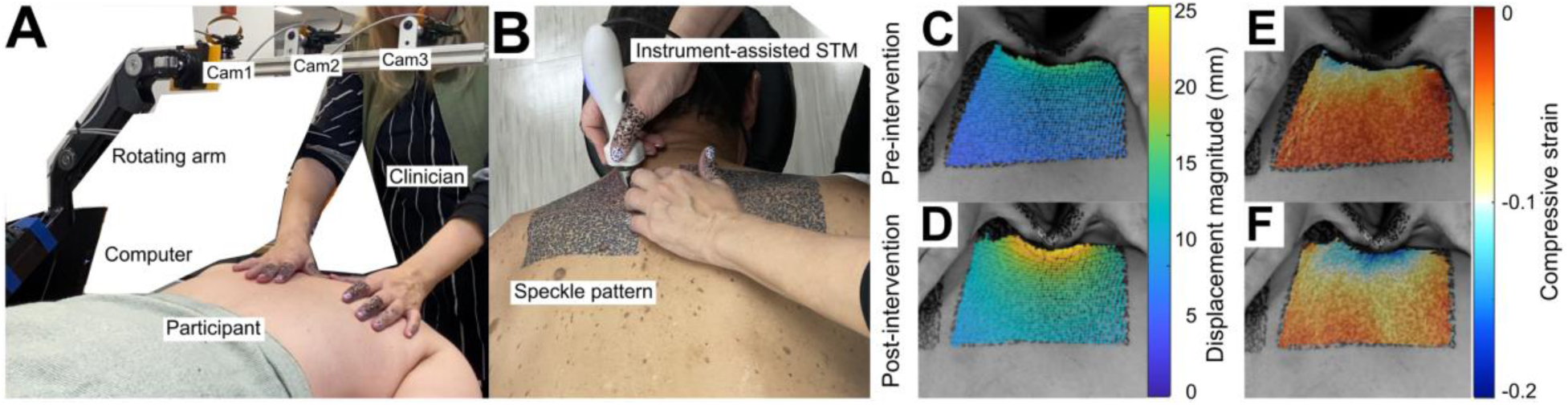
Equipment setup and STM intervention. (a) The clinician assesses the mobility of tissue of the right cervicothoracic region of a participant while the skin surface is imaged with three overhead cameras mounted to a rotating arm. (b) The clinician performs the instrument-assisted part of the STM intervention in the left cervicothoracic region. (c)-(f) Examples comparisons of tissue mobility pre- and post-intervention. (c)-(d) Colormaps overlaid on raw images showing change in skin surface displacement during a manual pull by clinician’s fingers in the superior direction towards the neck (c) before and (d) after an intervention. The blue to yellow color hues indicate greater skin mobility after the intervention. (e)-(f) Colormaps overlaid on raw images showing change in skin surface compressive strain over the same two manual pulls. The highest compressive strain (blue) is found near the area in contact with the clinician’s fingers and further from that point tends towards zero (red). More compressive strain is observed after the intervention compared to before the intervention for this participant

The manual STM phase began with effleurage, a series of gliding strokes with light pressure to warm the tissue before progressively increasing force. This was followed by cross-fiber strokes, applied perpendicularly to muscle fibers, and sweeping strokes, applied parallel to fiber alignment, to target different layers of fascia. Additional techniques, including petrissage (kneading motions to lift and roll soft tissues), skin rolling (mobilizing the skin and subcutaneous layers by lifting and rolling), and localized strumming strokes or sustained pressure (focused application of force to tender spots for up to one minute), were used to address deeper myofascial structures [57].

The IASTM portion employed a quantifiable soft tissue manipulation device (QSTM, Precision Care Technologies, Inc., Indianapolis, IN) [58, 59], using sweeping and strumming strokes to address localized tender or restricted tissues. Strokes were applied in multiple directions, including inferior-to-superior, superior-to-inferior, and curved medial-lateral patterns, with a stroke length of approximately 1 to 3 inches [60, 61]. The session concluded with effleurage strokes, reducing applied force and restoring baseline tissue tension in preparation for the post-intervention assessment. The average peak force applied during IASTM sweeping and strumming strokes was 12.12 ± 2.18 N (range: 9.78 - 16.12 N), calculated as the peak tri-axial resultant force for each stroke. The average stroke frequency was 1.46 ± 0.29 Hz, ranging from 0.96 to 1.86 Hz. The angle of application averaged 54.75 ± 20.15°, with a range of 40.5° to 69°.

### Equipment Setup and Participants

A portable massage table was equipped for participants to assume a prone position for STM assessment and intervention (Fig. 2(a)). The table’s face cradle was carefully adjusted to ensure the neck remained strain-free. All surfaces were sanitized regularly. Three cameras, mounted on a rotating arm affixed to the table, were positioned to capture images for the DIC analysis. Comprehensive details of this setup have been described previously [53].

In total, 19 participants were recruited (mean age 34.1 years, range 20–67 years; 9 males and 10 females). Participants were generally healthy adults but reported a range of cervicothoracic pain levels at baseline, including some with no pain (n=6) and others with pain consistent with benign cervicothoracic myofascial origin (n=13). All participants provided written consent, as approved by the University of Virginia Institutional Review Board of Social and Behavioral Sciences (Protocol #6201; Approved October 25th, 2023). This study was conducted as part of the clinical trial, “Optical Measurements of the Skin Surface to Infer Distinctions in Myofascial Tissue Stiffness (OptMeasSkin),” registered at ClinicalTrials.gov (ID: NCT06390085).

### Experimental Procedure

Approximately 24 hours before the scheduled session, participants completed consent forms and underwent the speckling procedure on both bilateral cervicothoracic regions (15 minutes). The study session began with a qualitative intake assessment (20 minutes), during which participants provided demographic and medical history information and reported their current pain in left and right cervicothoracic regions using a 0–10 numeric rating scale (0 = no pain, 10 = worst imaginable pain). The self-reported pain ratings were used as a proxy assumption for perceived tissue stiffness, rather than clinician assessment, to eliminate any bias that might be introduced by our study procedural constraints. The same clinician performed both STM stretch assessments and STM intervention for all applicable participants and was not blinded to intervention status (pre vs. post) or participants’ pain reports. The STM intervention is intentionally adaptive, informed by palpation and participant feedback, consistent with real-world clinical practice. The stretch assessment, by contrast, follows a fixed protocol (pull angle, ramp timing, trial count, anatomical reference points) established once per session, constraining the parameters through which awareness of pre/post status or pain reports could influence the assessment itself.

Following the intake assessment, participants were positioned on a massage table in a standardized prone position, with their arms at their sides (palms up) and feet resting on a bolster. The cervicothoracic junction (C7/T1), superior medial border of the scapulae, and distal insertion of the levator scapulae were palpated and marked with a black dot, bilaterally. For the pre-intervention assessment, STM stretch was applied in two directions (superior and inferior) first on the right side of the body, then on the left side (Fig. 2(c, e)). Each maneuver was repeated three times for a total of three trials per direction per body side for a total of 12 stretch assessments per participant pre-intervention, over a session duration of 20 minutes.

After the pre-intervention assessment, participants received a 5 minute break before undergoing the STM intervention protocol, which was performed bilaterally. A second 5 minute break was provided before the post-intervention STM assessment (Fig. 2(d, f)). After the post-intervention assessment, the clinician conducted a 10 minute qualitative outtake session, during which participants again reported their current pain in left and right cervicothoracic regions using the 0-10 numeric rating scale. Although both intake and outtake pain ratings were collected, only intake values were analyzed; outtake ratings were documented but not used as outcomes.

As the present study focuses on biomarker sensitivity rather than treatment efficacy, a typical standardized control or sham group was not included. Instead, two participants underwent repeated STM assessments separated by an equivalent time interval without intervention, providing a no-change condition for comparison.

### 3D Digital Image Correlation and Semi-Permanent Speckling Method

This study utilizes 3D digital image correlation (3D-DIC), a non-contact optical tracking technique, to measure fields of skin surface displacement and strain during soft tissue manipulation. DIC works by dividing an image into localized subsets, allowing the detailed analysis of movement and deformation through cross-correlation of pixel patterns from multiple stereo camera angles [62, 63]. Stereo camera calibration ensures accurate 3D reconstruction and enables the stitching of multiple surfaces together to avoid occlusion and account for highly curved regions [64].

To achieve high-resolution displacement fields for digital image correlation, a semi-permanent ink tattoo method was developed to create consistent, high-contrast speckle patterns. Four custom high-density speckle pattern stickers (∼1.5 mm speckle diameter; 15.2 cm x 7.6 cm; InkBox, Toronto, Canada) were applied bilaterally to the cervicothoracic region of each participant to create two regions of interest (15.2 cm x 15.2 cm; 231.0 cm²), one on the left side and one on the right side (Fig. 2(b)). The tattoo stickers remained in place for 1 hour, during which the ink reacted with the skin’s proteins to create a blue stain. After removal of the tattoos and over the course of 24 hours, the light blue speckles deepen to a dark blue, enhancing pattern-to-skin contrast for DIC tracking. The formula works on all skin tones, with darker skin pigmentations generating darker tattoo patterns. Patterns remain visible for up to ten days, after which the skin regenerates and the color naturally fades.

Data were collected using the open-source MultiDIC software [64], which builds on the Ncorr platform [65]. Synchronous video recording was performed with three cameras at 30 frames per second and 1920 by 1080 resolution (∼5 pixels/mm), which was later down-sampled to 15 frames per second for processing. Grayscale images were analyzed using subsets of 20 pixel radius and 10 pixel spacing, optimized for the speckle pattern and deformation characteristics. The DIC pipeline generated the 3D displacement and principal strain fields, which formed the basis for the eleven biomarkers.

Full details of the skin speckling procedure and DIC methodology can be found in prior work [53].

### Strain-Based Tissue Biomarkers

Eleven strain-based tissue biomarkers were developed to quantify soft tissue mobility and detect asymmetries within individuals across by sides, tissue pull directions, and pre- to post-intervention condition. Mobility is characterized from the spatial and temporal evolution of skin surface motion during clinician-applied pull, focusing on two key behaviors: tissue glide (how far tissue moves in response to loading) and tissue deformation (how much tissue stretches or compresses under that load). Restricted tissue typically behaves as if tethered, producing interrupted or discontinuous glide and, consequently, elevated deformation. After intervention, restrictions are expected to lessen, with smoother, more continuous movement to follow the direction of applied pull, reflected by greater glide and reduced deformation.

Of the eleven biomarkers, five primarily capture tissue glide and six describe tissue deformation. Tissue glide biomarkers include: *maximum pull (mm)*, the largest displacement near the point of force application; *far region displacement (mm)*, the largest displacement at the far edge of the tracked region; *pull propagation (%)*, the ratio of far to near displacement, where higher values indicate that more of the applied motion transmits across the region; *ramp-on pull velocity (mm/s)*, the initial rate of tissue displacement; and *ramp-on pull duration (s)*, the total time over which the tissue is actively pulled.

Tissue deformation biomarkers describe localized and global strain patterns. *Maximum tensile strain (unitless)* is the 95^th^ percentile of the firstprincipal Lagrangian strain field across the surface over time, representing peak tissue expansion, whereas *maximum compressive strain (unitless)* is the 5^th^ percentile of the second principal strain field, reflecting peak contraction. *Ramp-on strain gradient (1/mm)* quantifies how quickly tensile strain accumulates per millimeter of pull, capturing early resistance to deformation. *Tensile strain at 5 mm (unitless)* and *strain-to-pull ratio at maximum pull (1/mm)* standardize tissue response at fixed and peak loading, respectively. *Total gross deformation (mm)* integrates tensile strain over displacement to summarize cumulative deformation in a way that is largely time-independent, as velocity effects are expected to be minimal given highly consistent pull delivery by the clinician. Together, these biomarkers provide a structured, objective framework for quantifying soft tissue glide and deformation *in vivo*.

### Statistical Analysis

Linear mixed-effects (LME) models were used to evaluate systematic trends while accounting for individual variability and repeated measurements. Population-level effects were assessed using unpaired models, whereas within-participant effects were assessed using paired models. Model assumptions of linearity, homoscedasticity, and normality of residuals were checked using residual and Q-Q plots, and statistical significance was set at *α* = 0.01.

For individual-level mobility analyses, biomarker values were first averaged across three trials per pull, with percent differences (Eq. 1) then computed for three categories of A, B comparisons: directional (superior, inferior), bilateral (left, right), and intervention (pre, post). A 25% cutoff,

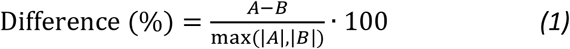

consistent with prior work [66–68], was used as a conservative threshold to distinguish physiological change from measurement variability. Biomarker comparisons with a precent difference magnitude exceeding this threshold were classified as different, and participants with at least 25% of comparisons exceeding the threshold were classified as exhibiting a pre-post intervention change, or a directional or bilateral asymmetry. The use of multiple comparisons to exceed threshold helps to minimize false positives and highlight consistent patterns of change. To evaluate the sensitivity of this threshold, the same classification procedure was also performed using 20% and 30% cutoffs. Exact (Clopper-Pearson) 95% confidence intervals (CI) [69] are reported for key summary proportions given the modest sample size.

Self-reported pain ratings collected at intake were used to characterize perceived bilateral asymmetries, with a ≥ 2 point difference on the 0–10 pain scale indicating a clinically meaningful discrepancy. Although subjective, such ratings have been associated with underlying tissue stiffness [70–72], supporting their use as a clinical reference in the absence of an objective gold standard. Intake pain ratings were therefore compared with DIC-derived mobility classifications, while outtake ratings were recorded but not analyzed to avoid bias from expectancy and immediate post-intervention influences.

## Results

### Participant Population Trends

Across all participants, linear mixed-effects models were used to assess mobility changes following intervention (pre vs. post), and asymmetries across pull directions (superior vs. inferior) and body sides (left vs. right). Two general types of analyses were conducted, first with all participants in aggregate and then within individual participants. Moreover, the analysis of the entire set of eleven strain-based biomarkers is provided in the Supplemental Material, whereas the Results section focuses on four selected biomarkers. The rationale for their selection is described at the end of the following section.

### Pre- to Post-Intervention Mobility Changes in Aggregate

With all participants, pull directions, and body sides aggregated, three biomarkers related to tissue glide (maximum pull, maximum far region displacement, and ramp-on pull velocity) and one biomarker related to tissue deformation (total gross deformation) increased significantly after the intervention (Fig. 3(a-d), Supp. Table S1). With regards to the first two tissue glide biomarkers, maximum pull increased by 3.4 mm (29.7 ± 9.7 mm pre compared to 33.1 ± 10.2 mm post; *t*(342) = 3.18, *p* < 0.01) (Fig. 3(a)) and maximum far region displacement rose by 2.7 mm (18.7 ± 9.3 mm pre compared to 21.4 ± 9.7 mm post; *t*(342) = 3.03, *p* < 0.01) (Fig. 3(b)). Although near and far region displacement both increased, pull propagation remained stable at approximately 65% (*t*(341) = 2.00, *p* > 0.01) (Supp. Fig. S1(c)), suggesting that the additional displacement was distributed proportionally across the tracked region. Moreover, pull velocity increased after the intervention, which suggests less resistance from the tissue during applied pull. In particular, ramp-on pull velocity climbed from 33.2 ± 16.0 mm/s to 42.1 ± 14.9 mm/s (Δ = 8.4 mm/s; *t*(341) = 4.29, *p* < 0.01) (Fig. 3(c)). With regards to the tissue deformation biomarker, total gross deformation was observed to increase (3.1 ± 1.7 mm to 3.7 ± 2.0 mm; *t*(342) = 3.35, *p* < 0.01) (Fig. 3(d)). Interestingly, although this biomarker is grouped with the deformation biomarkers, its increase can be primarily attributed to greater tissue displacement, as maximum tensile strain remained unchanged (*t*(342) = 0.99, *p* > 0.01) (Supp. Fig. S1(a)).

**Fig. 3.**
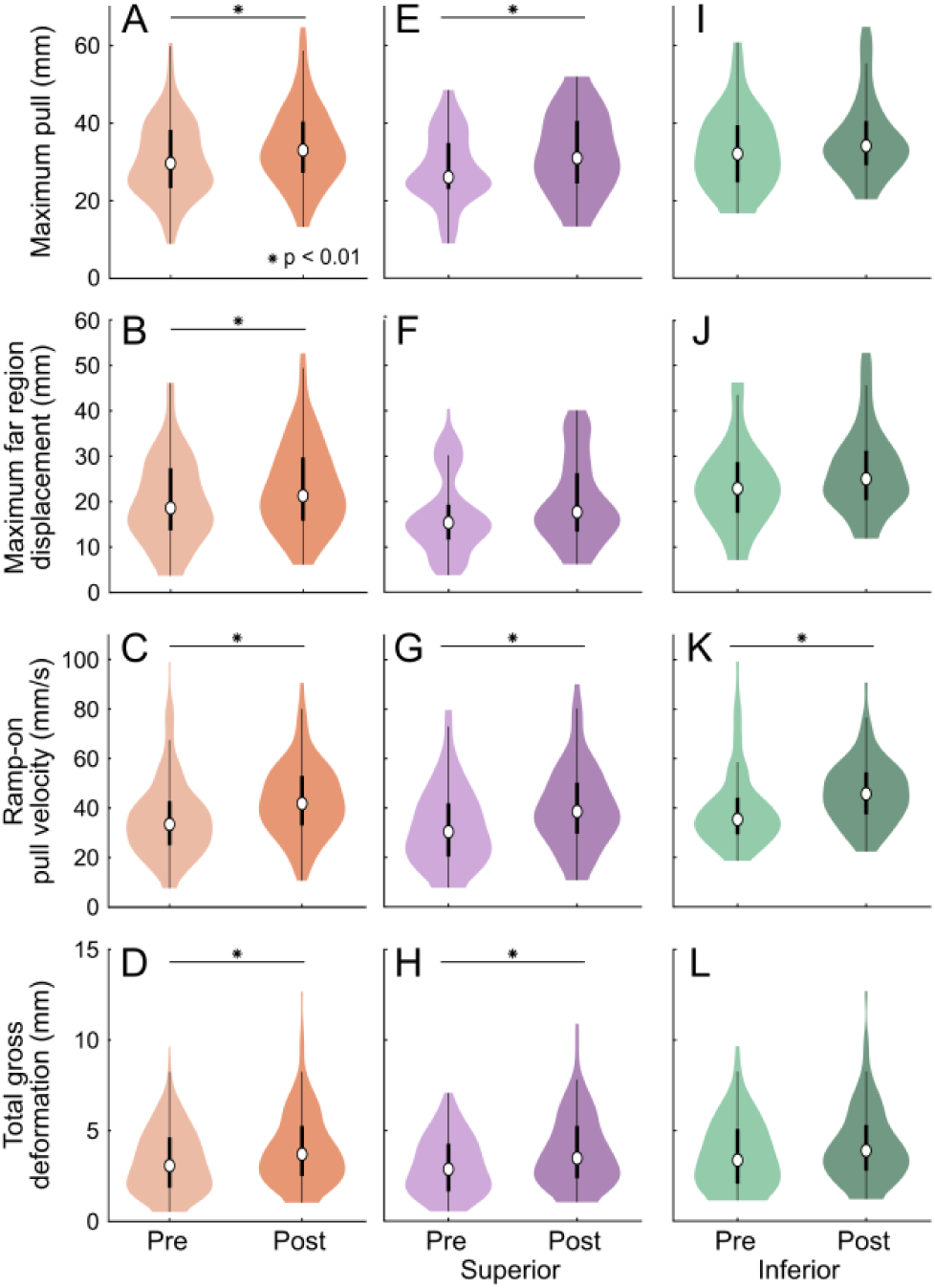
Participant population trends: pre- to post-intervention changes in mobility, presented both in aggregate and separated by pull direction (superior, inferior). (a)-(d) Distribution of four selected biomarkers across all participants, with body sides and pull directions aggregated, separated by intervention condition, and analyzed using linear mixed-effects (LME) models (α = 0.01). Post-intervention, in (a), the skin and subcutaneous tissue could be pulled 3.34 mm farther (29.74 ± 9.74 mm compared to 33.08 ± 10.16 mm, median ± SD) and in (b), reached 2.68 mm more maximum far region displacement. Similarly, in (c), ramp-on pull velocity increased 8.35 mm/s (33.72 ± 16.00 mm/s compared to 42.07 ± 14.89 mm/s) and in (d), total gross deformation increased 0.64 mm (3.11 ± 1.74 mm compared to 3.73 ± 2.04 mm). (e)-(h) Data isolated for the superior pull direction, separated by intervention condition. Significant changes were observed in four biomarkers (three shown): (e) maximum pull, (g) ramp-on pull velocity, and (h) total gross deformation increased post-intervention. (i)-(l) Data isolated for the inferior pull direction, separated by intervention condition. Significant changes were observed in one biomarker, (k) ramp-on pull velocity. Specifically, post-intervention, ramp-on pull velocity increased by 8.11 mm/s in the superior direction (30.55 ± 15.66 mm/s compared to 38.66 ± 16.38 mm/s) and 10.29 mm/s in the inferior direction (35.50 ± 15.71 mm/s compared to 45.79 ± 12.70 mm/s)

Soft tissue manipulation interventions aim to improve mobility by increasing tissue glide and reducing stiffness [73–76], yet no established mechanical standard defines what constitutes “more mobile” tissue. Of the entire set of eleven biomarkers (Supp. Fig. S1), the four biomarkers of focus in Figure 3 showed consistent, systematic changes following intervention—maximum pull, maximum far region displacement, ramp-on pull velocity, and total gross deformation. As all four biomarkers increased post-intervention, higher values correspond to greater mobility within each participant. Based on this analysis and that to be described below in the section *Biomarker Sensitivity*, we focus upon this data-driven subset of four selected biomarkers throughout the remainder of the manuscript. In particular, they are used to evaluate mobility relative to each individual, ensuring that within-participant classifications rely on intervention-responsive metrics. Moreover, this subset enables individual-level evaluation of (1) pre- to post-intervention mobility changes (whether a body side increased or decreased in mobility) and (2) bilateral mobility asymmetries at baseline (which body side was more mobile pre-intervention).

### Pre- to Post-Intervention Mobility Changes Separated by Pull Direction

With all participants and body sides aggregated, pre- to post-intervention mobility changes differed by pull direction. Specifically, ramp-on pull velocity increased following intervention in both pull directions, while only the superior direction exhibited accompanying changes in other mobility biomarkers, predominantly reflecting increased tissue deformation.

In the superior direction, four biomarkers (maximum pull, ramp-on pull velocity, total gross deformation – Fig. 3(e, g, h); and maximum compressive strain – Supp. Fig. S1(i)) changed systematically following intervention. Maximum pull increased by 4.9 mm (*t*(170) = 2.72, *p* < 0.01) (Fig. 3(e)); however, maximum far region displacement remained stable. Instead, maximum compressive strain increased in magnitude (*t*(170) = -3.10, *p* < 0.01) (Supp. Fig. S1(i)), indicating that additional loading was accommodated primarily through greater local deformation rather than extended tissue glide.

Across both pull directions, ramp-on pull velocity increased following intervention, by 8.1 mm/s superiorly (*t*(169) = 3.15, *p* < 0.01) and 14.4 mm/s inferiorly (*t*(170) = 3.06, *p* < 0.01) (Fig. 3(g, k)). As the pull maneuver is expertly modulated by tissue response through tactile feedback, a higher ramp-on pull velocity suggests reduced initial tissue resistance, allowing the same pull to be executed more rapidly before reaching a comparable end-feel. Taken together, these directional dependencies indicate that baseline anatomy and underlying structures shape how mobility changes are expressed. Inferior pulls, which exhibited greater baseline tissue glide [53], showed limited additional glide or deformation post-intervention, whereas superior pulls—where glide is more anatomically constrained—accommodated increased loading primarily through greater local tissue deformation.

### Post-Intervention Asymmetries in Tissue Mobility Given Pull in Superior vs. Inferior Direction

With all participants and body sides aggregated, post-intervention data showed consistent directional asymmetries. In particular, tissue glide biomarkers were higher for pulls in the inferior direction (Supp. Fig. S2(l-n, r)), whereas tissue deformation biomarkers were higher in magnitude for pulls in the superior direction (Supp. Fig. S2(q, s, u, v)), mirroring pre-intervention patterns [53]. Larger magnitudes of maximum pull (Δ = 3.4 mm; *t*(194) = 3.07, *p* < 0.01), maximum far region displacement (Δ = 7.4 mm; *t*(194) = 5.38, *p* < 0.01), ramp-on pull velocity (Δ = 9.5 mm/s; *t*(194) = 3.62, *p* < 0.01), and pull propagation (Δ = 11.0%; *t*(194) = 7.82, *p* < 0.01) indicated greater tissue glide in the inferior direction (Supp. Fig. S2(l–n,r)). In contrast, superior pulls exhibited higher maximum compressive strain (Δ = 0.037; *t*(194) = 7.69, *p* < 0.01), ramp-on strain gradient (Δ = -0.001; *t*(194) = -3.85, *p* < 0.01), strain at 5 mm (Δ = -0.017; *t*(194) = -4.50, *p* < 0.01), and strain-to-pull ratio (Δ = -0.001; *t*(194) = -3.97, *p* < 0.01), reflecting greater local deformation (Supp. Fig. S2(q,s,u,v)). These directional asymmetries are consistent with underlying anatomy: superior pulls engage the shoulder complex and overlapping musculature (trapezius, rhomboids, erector spinae), which restrict glide and promote deformation, whereas inferior pulls are applied over the more uniform latissimus dorsi, allowing motion to propagate along a long, continuous surface with greater “runway” for tissue glide.

### Post-Intervention Asymmetries in Tissue Mobility Given Assessment of Left vs. Right Body Side

With all participants and pull directions aggregated, post-intervention data showed no bilateral asymmetries (*p* > 0.01) (Supp. Fig. S2(w-g1)), in agreement with pre-intervention findings [53], and supporting the expectation of anatomical symmetry along the spine. This absence of population-level asymmetry reflects the fact that left-right mobility differences vary in direction and magnitude across individuals, such that aggregation obscures asymmetries that are only detectable through within-participant comparisons. Accordingly, these results motivate the individual-level analyses presented below, where bilateral mobility asymmetries are evaluated relative to each participant.

### Individual Trends

To capture changes that may be obscured by population-level statistics, we evaluated changes within individual participants. In particular, the four biomarkers defined in *Participant Population Trends—Pre- to Post-Intervention Mobility Changes in Aggregate* (maximum pull, maximum far region displacement, ramp-on pull velocity, and total gross deformation) were evaluated using the predefined thresholds (*Methods—Statistical Analysis*) to assess mobility changes following intervention (pre vs. post), and asymmetries in tissue mobility given pull direction (superior vs. inferior) and body side assessed (left vs. right).

### Pre- to Post-Intervention Mobility Changes

The results indicate that 16 of 17 treated participants (94.1%; 95% CI: 71.3-99.9%) changed in mobility after intervention, 15 of which (88.2%; 95% CI: 63.6-98.5%) demonstrated mobility improvements. The classification process is illustrated in Figure 4(a) for six pulls in the inferior direction on the left body side of a representative participant (P13). Following the STM intervention, the tissue allowed for a 33.2% increase in maximum pull (24.2 mm pre; 36.3 mm post) and a 47.0% increase in ramp-on pull velocity (25.0 mm/s pre; 47.2 mm/s post). Across the selected biomarkers (Fig. 3(a-d)), all four surpassed threshold given pulls in the inferior direction, while only one surpassed threshold given pulls in the superior direction. In total, five of eight biomarkers (62.5%) exceeded threshold across both body sides, meeting the criterion for a mobility change (Fig. 4(b); *Methods–Statistical Analysis*, *i.e.*, 25%). Similar analyses were performed across the cohort of 19 participants, with results reported in Table 1 and shown in Figure 4(c), separated by body side.

**Fig. 4.**
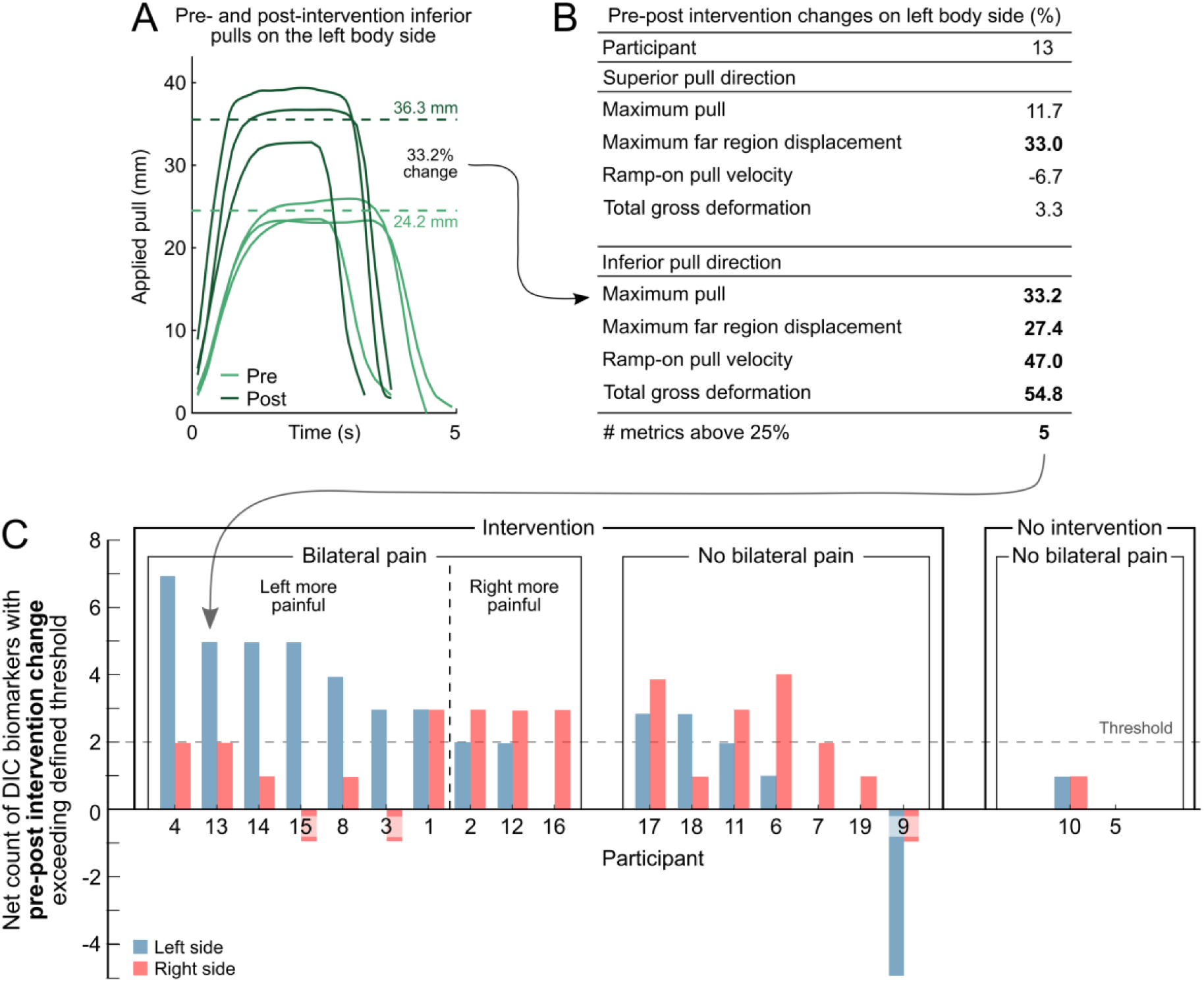
Individual trends: pre- to post-intervention mobility changes. (a) Time-series traces of a series of pulls in the inferior direction pre-intervention (light green) and post-intervention (dark green) on the left body side for a representative participant (P13). Post-intervention, the tissue was able to accommodate 12.1 mm more maximum pull (33.2% increase) at a 22.2 mm/s faster ramp-on pull velocity (47.0% increase). (b) Mobility changes were identified across both pull directions and body sides (only left shown), yielding eight total comparisons per participant. Biomarker changes were calculated as post-minus pre-intervention, such that positive values above threshold indicate greater mobility post-intervention and negative values above threshold indicate greater mobility pre-intervention. On the left body side, P13 exceeded the predefined threshold (*Methods— Statistical Analysis*) for one biomarker in the superior pull direction (25%) and in four biomarkers in the inferior pull direction (100%), for a total of five on the left body side. (c) Bars show, for each participant, the net number of biomarker comparisons exceeding threshold on left (blue) and right (red) body sides. A body side was classified as improved post-intervention if it exhibited a net increase of two or more comparisons exceeding threshold (positive change; dashed line) or as declined if it exhibited a net decrease of two or more comparisons exceeding threshold (negative change). Post-intervention, 15 treated participants (88.2%; P4, P13, P14, P15, P8, P3, P1, P2, P12, P16, P17, P18, P11, P6, P7) improved on at least one body side, while one treated participant (P9) declined on the left side. Participants are grouped by intervention status and by the presence of bilaterally asymmetrical pain at baseline, including which body side was reported as more painful. Among participants with bilaterally asymmetrical pain (n=10), nine (90%; P4, P13, P14, P15, P8, P3, P2, P12, P16) demonstrated greater mobility improvements on their more painful body side, while one (P1) showed equal improvements on both sides. For example, Participant 4 reported greater baseline pain on the left side, which improved post-intervention for seven biomarkers (87.5%), compared to two biomarkers (25%) on the right side. Two participants (P5, P10) did not receive an intervention and did not meet the criterion for a mobility change on either body side

**Table 1.** Individual trends: pre- to post-intervention mobility changes.

| Mobility changes between pre-intervention and post-intervention on the left body side (%) |  |  |  |  |  |  |  |  |  |  |  |  |  |  |  |  |  |  |  |
| --- | --- | --- | --- | --- | --- | --- | --- | --- | --- | --- | --- | --- | --- | --- | --- | --- | --- | --- | --- |
| Participant | 1 | 2 | 3 | 4 | 5 | 6 | 7 | 8 | 9 | 10 | 11 | 12 | 13 | 14 | 15 | 16 | 17 | 18 | 19 |
| Superior pull direction |  |  |  |  |  |  |  |  |  |  |  |  |  |  |  |  |  |  |  |
| Maximum pull | 1.6 | -1.0 | 10.6 | 21.0 | 0.5 | 24.3 | 14.4 | 39.5 | -18.0 | 6.3 | 9.6 | 21.5 | 11.7 | 21.6 | 28.4 | -11.5 | 5.3 | 7.6 | 2.9 |
| Far region displacement | 4.8 | -0.4 | 18.2 | 46.2 | 4.2 | 21.0 | 24.6 | 63.6 | -20.6 | 17.0 | 26.3 | 21.4 | 33.0 | 16.0 | 34.1 | -21.6 | 11.7 | 8.4 | 4.5 |
| Ramp-on pull velocity | 63.8 | 60.1 | 25.8 | 67.3 | 10.9 | -4.8 | 24.3 | 11.5 | -27.6 | 5.3 | 37.8 | 38.8 | -6.7 | 29.9 | 51.5 | 8.8 | 3.2 | 15.3 | 16.1 |
| Total gross deformation | 37.8 | 30.4 | -9.2 | 68.6 | -7.0 | 41.8 | 14.6 | 47.8 | -29.3 | 6.0 | 11.6 | 47.0 | 3.3 | 38.7 | 39.5 | -2.7 | 23.1 | 14.4 | 17.9 |
| Inferior pull direction |  |  |  |  |  |  |  |  |  |  |  |  |  |  |  |  |  |  |  |
| Maximum pull | 16.3 | -7.6 | 6.0 | 34.3 | -0.5 | -9.2 | -5.8 | 17.0 | -21.1 | 18.0 | -1.8 | -10.7 | 33.2 | 24.3 | 22.0 | -4.3 | 24.8 | 19.6 | 12.0 |
| Far region displacement | 12.8 | 8.0 | 4.6 | 36.8 | -1.5 | -2.7 | -11.6 | 39.5 | -25.8 | 18.0 | 1.5 | 7.8 | 27.4 | 26.5 | 24.2 | -2.7 | 29.2 | 30.7 | 12.5 |
| Ramp-on pull velocity | 65.7 | 51.2 | 45.8 | 52.8 | 22.9 | -12.1 | -14.6 | 2.7 | -36.0 | 17.2 | 18.2 | 6.6 | 47.0 | 43.4 | 12.7 | 15.0 | 31.1 | 40.3 | -10.1 |
| Total gross deformation | 18.1 | -25.0 | 38.1 | 39.2 | -6.9 | -22.9 | -19.5 | 3.4 | -41.9 | 34.3 | 1.0 | -24.3 | 54.8 | 61.3 | 36.1 | -3.6 | 44.1 | 32.3 | 21.5 |
| Left total | 3 | 2 | 3 | 7 | 0 | 1 | 0 | 4 | -5 | 1 | 2 | 2 | 5 | 5 | 5 | 0 | 3 | 3 | 0 |
| Mobility changes between pre-intervention and post-intervention on the right body side (%) |  |  |  |  |  |  |  |  |  |  |  |  |  |  |  |  |  |  |  |
| Participant | 1 | 2 | 3 | 4 | 5 | 6 | 7 | 8 | 9 | 10 | 11 | 12 | 13 | 14 | 15 | 16 | 17 | 18 | 19 |
| Superior pull direction |  |  |  |  |  |  |  |  |  |  |  |  |  |  |  |  |  |  |  |
| Maximum pull | 7.1 | 6.5 | 3.9 | 8.6 | 3.0 | 25.3 | 21.8 | 3.0 | 5.6 | -2.2 | 27.2 | 30.5 | 8.4 | 11.6 | 5.1 | -3.2 | 22.2 | -8.5 | 11.3 |
| Far region displacement | 5.3 | 4.5 | 4.6 | 5.3 | 0.7 | 12.3 | 25.0 | 7.0 | 5.5 | -1.0 | 47.3 | 40.7 | 3.4 | 9.5 | 14.0 | -13.9 | 22.0 | -9.3 | 16.3 |
| Ramp-on pull velocity | 60.1 | 58.0 | 9.1 | 36.9 | 13.8 | 43.7 | 22.0 | 3.2 | -3.4 | -6.2 | 17.7 | 20.1 | 22.4 | 31.7 | -5.1 | 11.5 | 11.7 | 6.4 | 29.6 |
| Total gross deformation | 39.6 | 34.8 | -30.6 | 1.9 | -6.2 | 42.6 | 28.4 | 3.5 | -1.6 | -5.7 | 31.5 | 39.0 | 31.1 | 24.5 | -7.5 | 27.9 | 40.5 | -33.1 | 9.0 |
| Inferior pull direction |  |  |  |  |  |  |  |  |  |  |  |  |  |  |  |  |  |  |  |
| Maximum pull | 2.9 | 13.4 | -20.1 | 2.9 | -4.4 | -0.1 | -7.8 | 20.5 | -15.8 | 15.9 | 14.8 | 6.4 | -17.3 | 1.8 | 10.6 | 17.7 | 47.2 | 23.5 | 2.8 |
| Far region displacement | -7.0 | 14.6 | 1.6 | 4.1 | 3.5 | 12.0 | -7.3 | 38.3 | -20.9 | 17.7 | 20.9 | 10.9 | -6.1 | 3.4 | 17.7 | 26.9 | 52.6 | 20.1 | 5.0 |
| Ramp-on pull velocity | 31.3 | 60.1 | 39.1 | 41.0 | 22.2 | 28.2 | 27.5 | -6.3 | -21.8 | 18.5 | -0.7 | 5.5 | 26.5 | 23.6 | -25.8 | 21.5 | 18.4 | 25.1 | 18.5 |
| Total gross deformation | -4.1 | 19.7 | -36.7 | 16.8 | -12.5 | -6.9 | -20.6 | 17.0 | -37.9 | 32.2 | 19.8 | 7.4 | -15.2 | 21.4 | 10.1 | 34.7 | 64.3 | 38.5 | 23.2 |
| Right total | 3 | 3 | -1 | 2 | 0 | 4 | 2 | 1 | -1 | 1 | 3 | 3 | 2 | 1 | -1 | 3 | 4 | 1 | 1 |

Post-intervention mobility changes were identified using the four selected biomarkers applied across both pull directions and body sides. Biomarker changes were computed as post- minus pre-intervention, such that positive values indicate greater mobility post-intervention and negative values indicate greater mobility pre-intervention (*Methods—Statistical Analysis*). The table reports, for each participant, pull direction, and body side, the number of selected biomarker comparisons exceeding the predefined threshold. Values are separated by pull direction (superior—top block; inferior—bottom block) and body side (left—top table; right—bottom table), with counts summed across directions to yield eight total comparisons per body side (four biomarkers × two directions). Comparisons exceeding threshold are bolded, with blue shading indicating positive values and red shading indicating negative values. A body side was classified as exhibiting post-intervention mobility improvement if it showed a net increase of two or more comparisons exceeding threshold (positive change, blue), or as declined if it showed a net decrease of two or more comparisons exceeding threshold (negative change, red). Post-intervention, 15 treated participants (88.2%; P1-P4, P6-P8, P11-P18) improved on at least one body side, while one treated participant (P9) declined on the left side. Two untreated participants (P5, P10) did not meet the criterion for a mobility change on either body side.

The results indicate that all ten participants with bilateral pain asymmetries improved on at least one body side, nine of whom showed relatively greater improvements on their more painful side (Fig. 4(c), Table 1). For example, Participant 13’s left side was reported as the more painful side at baseline (pre-intervention) and improved in 5 biomarkers (62.5%) post-intervention, compared to 2 on the right side (25%). Of the seven participants who received treatment but reported no bilateral pain asymmetries, five showed improvements on at least one body side that exceeded the threshold, while one participant (P9) showed a decline in mobility on their left side—the only case of reduced mobility in the population. Of the two participants who did not receive an STM intervention (P5 and P10), neither showed a change in mobility above threshold (Fig. 4(c)).

This classification was robust to the choice of threshold: reclassifying participants using 20% and 30% cutoffs in place of 25% yielded consistent results in direction and magnitude (Supp. Table S3), with the proportion of treated participants classified as improved ranging from 76.5% to 94.1%.

### Pre-Intervention Asymmetries in Tissue Mobility Given Assessment of Left vs. Right Body Side

Pre-intervention, ten participants (52.6%) exhibited bilateral mobility asymmetries at baseline, of which seven (P4, P13, P1, P14, P8, P15, P10) were classified as more mobile on their right body side and three (P2, P16, P6) were classified as more mobile on their left (Fig. 5, Supp. Table S2). Asymmetries were identified using the four selected biomarkers applied across both pull directions (left superior vs. right superior; left inferior vs. right inferior), yielding eight comparisons per participant. Biomarker comparisons were calculated as right minus left side, such that positive values indicate greater mobility on the right side and negative values indicate greater mobility on the left (*Methods—Statistical Analysis*). A body side was classified as more mobile only if it exceeded the contralateral side by two or more comparisons.

**Fig. 5.**
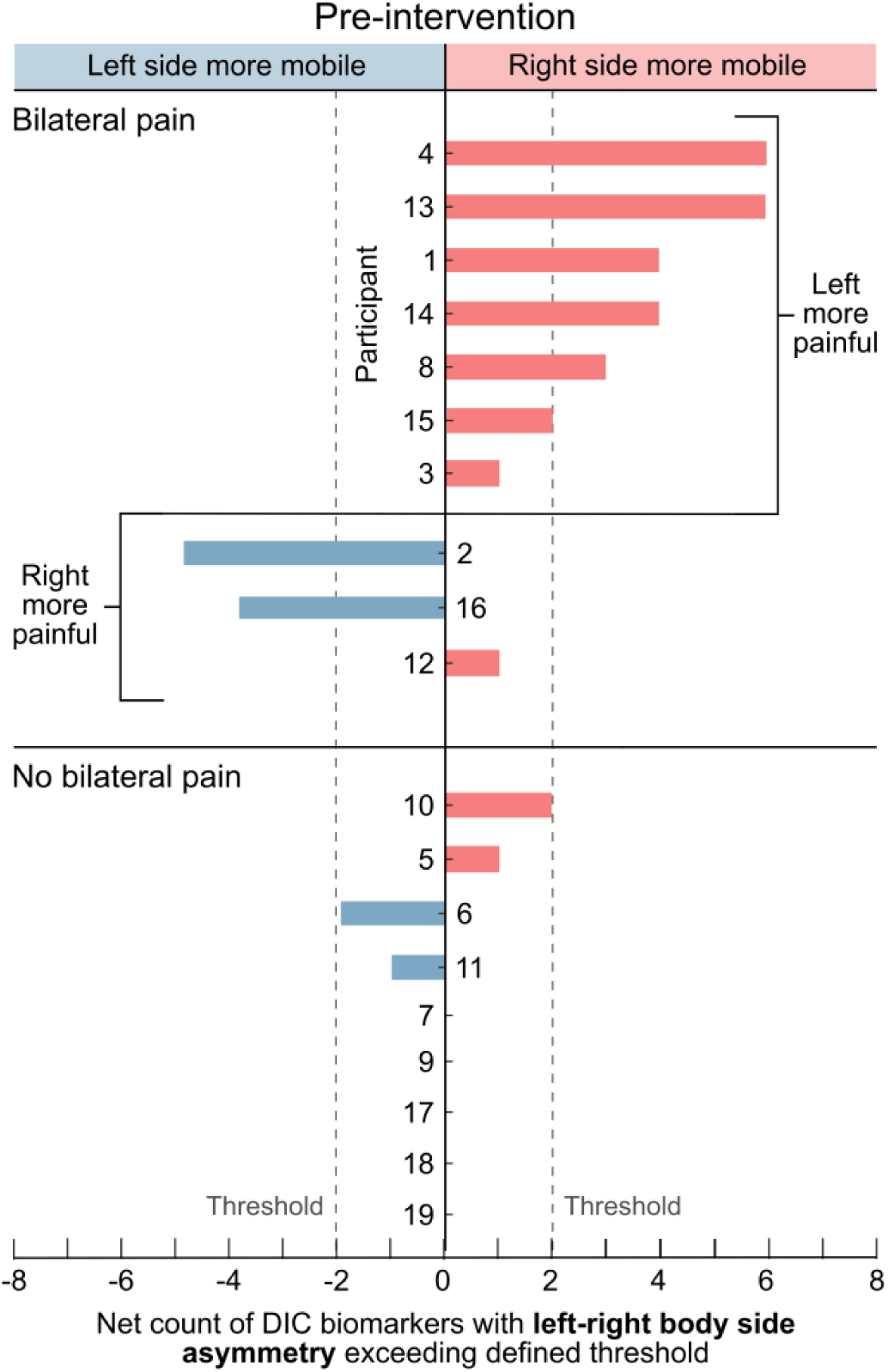
Individual trends: pre-intervention asymmetries in tissue mobility given assessment of left vs. right body side. Asymmetries in bilateral mobility were identified using the four selected biomarkers applied to pre-intervention data across both pull directions (left superior vs. right superior; left inferior vs. right inferior), yielding eight comparisons per participant. Biomarker comparisons were calculated as right minus left side, such that positive values indicate greater mobility on the right side and negative values indicate greater mobility on the left. Each comparison was counted toward the body side with the larger value if it exceeded the predefined threshold (*Methods—Statistical Analysis*). Bars show, for each participant, the net number of comparisons exceeding threshold. A body side was classified as more mobile only if it exceeded the contralateral side by two or more comparisons (dashed line). Ten participants (52.6%) exhibited bilaterally asymmetrical mobility at baseline with seven (P4, P13, P1, P14, P8, P15, P10) classified as more mobile on the right body side and three (P2, P16, P6) as more mobile on the left. Eight of these participants (80%) also reported bilaterally asymmetrical pain during intake. In all eight cases, the more mobile side corresponded to the less painful side: six right-more-mobile participants (85.7%; P4, P13, P1, P14, P8, P15) reported greater pain on the left, while two left-more-mobile participants (66.7%; P2, P16) reported greater pain on the right. Two participants (P3, P12) reported bilaterally asymmetrical pain but did not meet the criterion for bilaterally asymmetrical mobility, while two participants (P10, P6) met the criterion for bilaterally asymmetrical mobility without reporting bilaterally asymmetrical pain

For example, Participant 4 had six biomarker comparisons (superior: 2; inferior: 4) that exceeded the predefined threshold and were larger on the right body side, resulting in a classification of greater pre-intervention mobility on the right side compared to the left side (Supp. Table S2; bolded). In contrast, although Participant 3 had two biomarker comparisons (superior: 1; inferior: 1) that exceeded the threshold and were larger on the right body side, they also had one comparison in the superior direction that exceeded the threshold and was larger on the left side.

As a result, neither body side exceeded the contralateral side by two or more comparisons, and Participant 3 was not classified as having bilaterally asymmetrical mobility at baseline.

Eight of the participants with bilaterally asymmetrical mobility pre-intervention (80%) also reported bilaterally asymmetrical pain during intake. In all eight cases, the participant’s more mobile side aligned with their less painful side, that is six right-more-mobile participants (P4, P13, P1, P14, P8, P15; 85.7%) reported more pain on their left sides, while two left-more-mobile participants (P2, P16; 66.7%) reported more pain on their right sides. Two participants (P3 and P12) reported bilaterally asymmetrical pain but were not found to be bilaterally asymmetrical in mobility. Similarly, two participants (P10 and P6) with bilaterally asymmetrical mobility reported no bilaterally asymmetrical pain, reinforcing that clinically relevant mobility asymmetries can persist even when pain perception appears balanced.

### Biomarker Sensitivity

Biomarker sensitivity was evaluated across the 19 participants and three comparison categories (directional, bilateral, pre-post). For each of the four selected biomarkers, the number of unique participants exhibiting at least one detection (25%) per category was counted (Fig. 6).

**Fig. 6.**
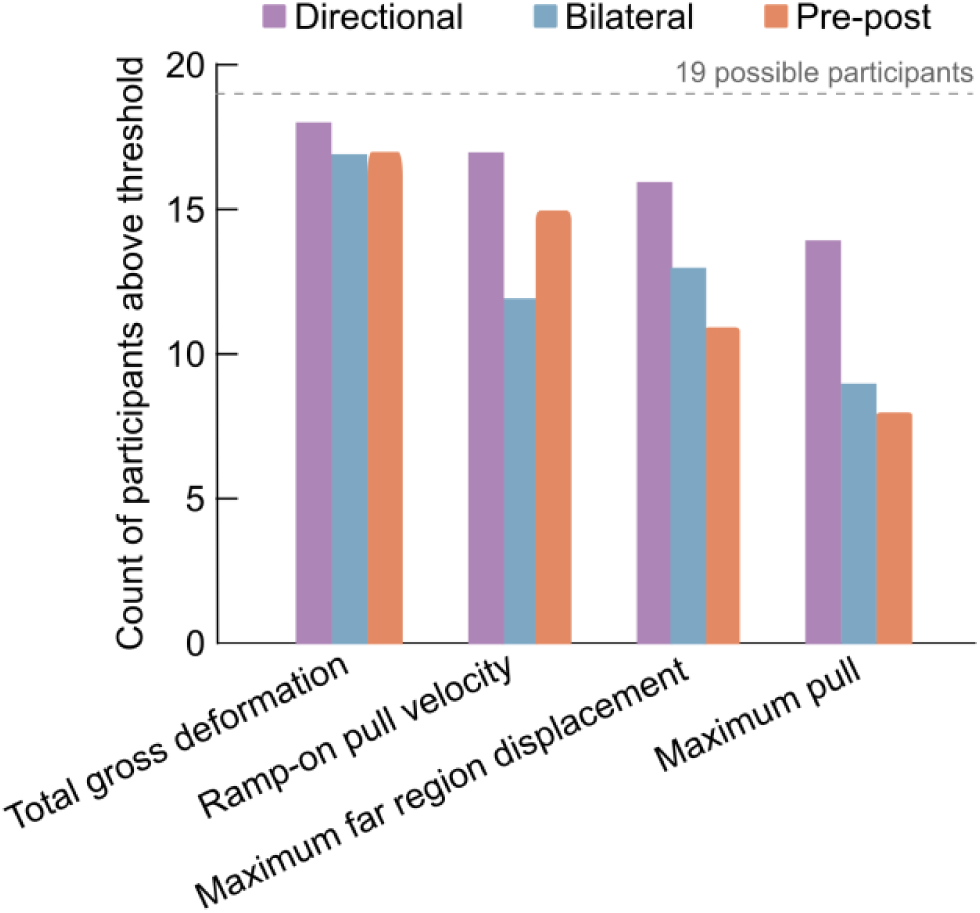
Biomarker sensitivity. Bars indicate the number of participants (out of 19) with at least one threshold-exceeding detection for each of the four selected biomarkers, separated by category (directional—purple, bilateral—blue, pre-post— orange). Total gross deformation and ramp-on pull velocity identified the most participants, with directional detections in 18 and 17 participants, bilateral in 17 and 12, and pre-post in 17 and 15, respectively. Maximum far region displacement and maximum pull identified fewer participants overall, particularly in the pre-post category (11 and 8 participants, respectively)

Total gross deformation exceeded threshold in the most participants overall, capturing all 19 participants at least once across the three categories. Within individual categories, it identified 18 participants in directional comparisons, 17 in bilateral, and 17 in pre-post. Ramp-on pull velocity identified 18 of 19 participants (94.7%) at least once, with strong representation across directional (17 participants), bilateral (12 participants), and pre-post (15 participants) categories. Maximum far region displacement also identified 18 participants overall, with detections concentrated in directional (16 participants) and bilateral (13 participants) categories, while pre-post analyses captured only 11 participants. Maximum pull identified the fewest participants overall (17 total), with 14 in directional, 9 in bilateral, and only 8 in pre-post categories. These results demonstrate that total gross deformation and ramp-on pull velocity provide the most robust participant coverage, while maximum pull shows more limited sensitivity, particularly for detecting pre-to-post intervention-related change.

Across all four biomarkers, the directional category consistently exceeded threshold in the most participants, capturing 85.5% of possible participant-comparison instances (65 of 76 total), compared with 67.1% for both bilateral and pre-post categories (Supp. Fig. S3). This pattern indicates that pull direction and anatomical orientation exert a dominant influence on observed soft tissue mobility, but that pre-post comparisons are frequently observed. The consistent dominance of directional comparisons across all biomarkers confirms that mechanical loading direction is a primary driver of strain-based mobility differences.

## Discussion

This study evaluated whether objective measures of tissue glide and deformation, optically derived from the skin surface using digital image correlation, could detect changes in soft tissue mobility immediately following STM intervention. Soft tissue manipulation is widely used to treat myofascial pain, but its effects on tissue mobility are typically inferred from subjective assessments. We tested two hypotheses: (1) that strain-based biomarkers at the skin surface would be sufficiently sensitive to detect changes in tissue mobility immediately before and after STM intervention, and (2) that tissue mobility would change more on the painful side for participants with bilaterally asymmetric pain. Our results support both hypotheses. Following STM intervention, four biomarkers changed systematically, with measurable increases in mobility on at least one body side for 88% of treated participants (15 of 17), and with greater increases in mobility on the more painful side for 90% of participants with bilaterally asymmetric pain (9 of 10).

### Biomarkers Can Detect Changes in Mobility Following STM Intervention

Consistent with the first hypothesis, the strain-based biomarkers detected STM-induced changes in tissue mobility both at the aggregate level and within individual participants. At the aggregate level, four biomarkers—maximum pull, maximum far region displacement, ramp-on pull velocity, and total gross deformation—changed systematically following STM intervention (Fig. 3(a-d)). Following intervention, the skin surface and underlying soft tissue system could be displaced laterally to a greater extent and could be loaded more rapidly; for example, maximum pull increased by 3.4 mm and ramp-on pull velocity by 8.4 mm/s. At the participant level, 94.1% of treated participants showed measurable changes on at least one body side (Fig. 4, Table 1).

Although the absolute magnitude of the changes in displacement may appear modest at 3.4 mm, similar small-scale alterations have been linked to meaningful changes in joint and soft tissue mechanics. For example, sub-millimeter increases in cervical facet joint gap (0.9 ± 0.40 mm) during spinal manipulation have been associated with measurable gains in intervertebral range of motion (ROM) and pain reduction in patients with neck pain [77]. The magnitude and interpretation of the present changes may depend on the mechanical properties of the tissue layers engaged by the clinician’s pull. Our DIC biomarkers quantify skin surface displacement and strain and therefore represent a composite response of the skin, subcutaneous tissue, superficial and deep fascia, interlayer glide planes, and deeper connective and muscular tissues. They do not directly measure fascial deformation, fascial gliding, or intrinsic tissue material properties. Prior ultrasound studies support the biological relevance of thoracolumbar connective tissue morphology and interlayer mechanics in musculoskeletal pain. In individuals with chronic low back pain relative to asymptomatic controls, prior studies have reported greater perimuscular connective tissue thickness and echogenicity [78] and approximately 20% lower thoracolumbar fascial shear strain during passive trunk flexion [79]. In contrast, a more recent ultrasound elastography study found greater lateral shear strain measures in individuals with nonspecific low back pain despite no significant group difference in thoracolumbar fascia thickness [80]. Together, these findings suggest that the relationships among fascial thickness, tissue composition, and mechanical glide behavior remain incompletely understood.

The observed increases in maximum pull and ramp-on pull velocity are consistent with clinicians’ use of manual techniques that involve repeated shear, compression, and distraction. Such interventions are thought to reduce resistance to inter-layer motion by altering the connective tissue matrix—through mechanisms including disruptions of collagen crosslinks, reduction of fascial adhesions, and reorganization of the extracellular matrix (ECM)—thereby lowering the effective viscosity opposing tissue sliding [81–84]. Regions requiring greater physiological interlayer sliding also contain higher concentrations of hyaluronan (HA), with HA content per gram of fascial tissue reported to range from approximately ∼6 µg in adherent fascia to ∼90 µg in highly mobile retinacula [85], which may support tissue lubrication and glide. Because soft tissues exhibit viscoelastic behavior, a reduction in rate-dependent resistance could permit tissue to be loaded more rapidly before comparable resistance is encountered, consistent with the observed increase in ramp-on pull velocity (Fig. 3(c)) [86]. Prior studies of STM and manual therapy likewise report localized changes in superficial and deeper tissue mechanics, including altered passive stiffness, ROM, fiber organization, and perfusion [81, 87–89].

### Bilaterally Asymmetric Mobility Aligns with Bilaterally Asymmetric Pain

Evidence was also observed to support the second hypothesis, *i.e.*, that participants with bilaterally asymmetric pain would exhibit larger mobility gains on their more painful side. Consistent with this hypothesis, the post-intervention results demonstrate that bilateral asymmetries in tissue mobility are not only detectable but can be improved using a brief STM intervention, at least in the short term. Overall, 88% of treated participants showed improved mobility on at least one body side, including all ten with bilaterally asymmetric pain at baseline. Nine (90%) of these exhibited greater mobility gains on their more painful body side (Fig. 4, Table 1). This side-specific improvement is compatible with experimental evidence that targeted soft tissue mobilization can induce quantifiable, localized changes in tissue mechanical behavior, including approximately 30-40% increases in strength and stiffness metrics in treated ligaments and tendons relative to contralateral injured, untreated tissue during early healing [83, 90]. Human studies of instrument-assisted soft tissue techniques similarly report measurable reductions in passive tissue stiffness and increases in joint range of motion localized to the treated region [91–94].

Prior to treatment, the biomarkers identified bilateral mobility asymmetries in ten participants, eight (80%) of whom also reported bilaterally asymmetric pain. In all eight cases, the more mobile side corresponded to the less painful side (Fig. 5, Supp. Table S2), in alignment with prior work linking lower pressure pain thresholds (PPT) to increased stiffness, reduced ROM, and greater symptom severity in myofascial pain [95–97]. Lower maximum pull and ramp-on pull velocity on more painful sides align with mechanisms of fascial densification—including collagen cross-linking, reduced hydration, and impaired interlayer glide—that increase stiffness and nociceptive drive while constraining motion [5, 78, 98–100]. These patterns correspond with clinical descriptions of myofascial trigger points, where localized stiffness and altered ECM organization are linked to pain and movement restriction [88, 101].

Together, these findings show that strain-based biomarkers can identify mechanically restricted, painful tissue and quantify its preferential improvement with treatment, positioning them as practical tools for linking local symptoms to measurable changes in soft tissue mobility.

### Need for Objective Metrics to Evaluate Effects of Soft Tissue Manipulation

Despite its widespread clinical use, STM assessments and interventions are typically evaluated using subjective outcomes such as pain ratings, ROM, or task-based measures like grip strength [74, 102–107]. While informative, these measures are influenced by a clinician’s perception, motivation, and testing context, and do not directly quantify local tissue properties at the treatment site. On the other hand, objective tools such as myotonometry and shear wave elastography (SWE) provide stiffness-related metrics but face limitations including probe orientation dependence, tissue anisotropy, and operator variability [108–111], which can contribute to inconsistent post-intervention findings. For example, SWE studies have shown that brief massage can produce an immediate but modest reduction in localized muscle stiffness (*e.g.*, ∼5% decrease in gastrocnemius shear modulus), although these effects return toward baseline within minutes [112]. More broadly, because these methods primarily quantify elastic properties under simplified compressive loading—rather than interlayer glide or deformation during functional, clinician-applied maneuvers—they may fail to capture dynamic lateral tissue mobility changes that persist beyond short-lived stiffness effects.

By contrast, the optical strain-based biomarkers introduced here quantify skin surface motion during hands-on clinical maneuvers without perturbing clinician-patient interaction, providing non-invasive, region-specific measures of soft tissue mobility. These metrics are not intended to replace self-reported outcomes but to provide objective mechanical context.

For most of the participants herein, the measured mobility and pain findings aligned, strengthening the inference that mechanical restriction contributes to symptoms. However, one participant (P9; Fig. 4) showed no pre-treatment mobility or pain asymmetry by our measures yet exhibited declining mobility on the left side following intervention. This divergence highlights that NRS pain scores, while useful, are subjective and multifactorial and may not fully capture psychophysiological contributors to mobility changes. One possibility is a localized increase in muscle tone driven by psychological stress during the STM session. Needle electromyography studies have shown that, in individuals with myofascial trigger points, these sites exhibit increased electrical activity during psychological stressors while adjacent non-tender muscle remains electrically silent, suggesting a mechanism by which emotional factors can modulate localized muscle tension independent of physical pain [113]. This case underscores the importance of combining objective biomarkers with clinical judgement and self-reported outcomes, as divergence may point to non-mechanical contributors, reporting variability, or temporal offsets between mechanical and symptomatic change [114]. Integrating these biomarkers with standard clinical assessments could enable more precise identification of mechanically restricted regions, support data-driven selection of STM techniques, and provide reproducible, tissue-level evidence of treatment effect that can be tracked across sessions and studies.

### Methodological Considerations and Work Needed for Clinical Translation

This study employed an observational framework in which an experienced clinician performed STM without imposed constraints, prioritizing ecological validity and reflecting real-world practice. Rather than estimating absolute forces or ground-truth tissue properties, the aim was to detect relative changes in tissue behavior and relate these patterns to self-reported pain. As this aspect of the study was exploratory, the small no-intervention case study (n=2) was not powered to rule out time-related or nonspecific effects of hands-on contact, but rather to provide a preliminary check that biomarker values did not notably change in the absence of intervention. A sham-controlled design is a planned next step to isolate the intervention-specific effect from these confounds. The modest sample size and single clinician are consistent with prior exploratory manual therapy studies but limit generalizability and preclude evaluation of inter-clinician variability [71, 110, 115]. Given these constraints, the reported percentages (*e.g.,* 88% and 90% improvement rates) should be interpreted as descriptive of this cohort rather than precise population estimates.

Additionally, mobility was assessed only immediately before and after intervention. As such, the durability of measured changes beyond this window is unknown, and longer-term follow-up is needed to determine whether observed gains persist. Notably, the semi-permanent speckle pattern used in this study remains visible for up to ten days, enabling repeated DIC assessment of the same tracked region, which we plan to leverage in future work to examine both the longitudinal effects of a single STM treatment and the cumulative effects of multiple treatments delivered within this window. Based on the promising findings of this initial study, subsequent studies should also expand to a larger, more diverse cohort and additional anatomical regions to examine how key demographic and clinical variables (*e.g.,* age, sex, BMI, condition type and chronicity) and anatomical site influence biomarker performance. Future work will also incorporate a blinded, two-clinician design, in which one clinician delivers the intervention and collects participant pain ratings while a second, blinded clinician performs the STM stretch assessment, to help quantify technique variability and validate whether the biomarker-based findings generalize across practitioners [116, 117].

Moreover, not all of the strain-based biomarkers may be necessary to achieve clinical adoption. In this study, we began with eleven biomarkers but down sampled those to a selected set of four. Two of those, total gross deformation and ramp-on pull velocity emerged as the most consistent and responsive indicators (Fig. 6), suggesting a compact subset may preserve diagnostic sensitivity while simplifying implementation. Presently, DIC processing remains time-intensive, requiring approximately one hour per trial to output biomarkers from raw video data. Reducing this complexity is an active goal. Future work will explore whether fewer frames (*e.g.,* static images at key points rather than continuous video) or reduced spatial resolution can preserve diagnostic sensitivity while easing computational burden. We believe it is first necessary to leverage the full spatiotemporal resolution of DIC to identify which features are most informative, before simplifying acquisition in ways that could sacrifice important information. In this sense, the current pipeline serves as a discovery tool for establishing what skin surface deformation can reveal about soft tissue mobility in musculoskeletal pain, with the longer-term goal of translating these insights into simpler, more scalable measurement approaches. Clinical translation will also require linking biomarker outputs to familiar clinical constructs (*e.g.,* ROM, perceived stiffness) and establishing thresholds for meaningful change at both population and individual levels.

## Data Availability

The data that support the findings of this study are available from the corresponding author (G.J.G.) upon reasonable request.

## Statements and Declarations

No conflicts of interest, financial or otherwise, are declared by the authors. A preprint of this manuscript has been posted on medRxiv (DOI: 10.1101/2025.07.31.25332529).

## Acknowledgements

This work was supported in part by the U.S. National Institutes of Health under Grants R01AT013186, R21AT011980, U24AT011969, and U24AT011970.

## Supplementary Information

Supplementary Figures S1-S3 and Supplementary Tables S1-S3 can be found in the file *Supplementary_Material.pdf*.

## Authors’ Contributions

A.R.K, M.T.L, and G.J.G conceived and designed research; A.R.K and M.T.L performed experiments; A.R.K. analyzed data; A.R.K, M.T.L, and G.J.G interpreted results of experiments; A.R.K prepared figures and drafted manuscript; A.R.K, M.T.L, and G.J.G edited and revised manuscript and approved final version of manuscript.

## Declarations

### Ethical Approval

All participants provided written consent, as approved by the University of Virginia Institutional Review Board of Social and Behavioral Sciences (Protocol #6201; Approved October 25th, 2023). This study was conducted as part of the clinical trial, “Optical Measurements of the Skin Surface to Infer Distinctions in Myofascial Tissue Stiffness (OptMeasSkin),” registered at ClinicalTrials.gov (ID: NCT06390085).

## References

1. Lucas, J. W., and I. Sohi. Chronic Pain and High-impact Chronic Pain Among U.S. Adults, 2023. Hyattsville, MD: National Center for Health Statistics (U.S.), 2024. 10.15620/cdc/169630

2. Nahin, R. L., A. Rhee, and B. Stussman. Use of Complementary Health Approaches Overall and for Pain Management by US Adults. JAMA 331:613–615, 2024. 10.1001/jama.2023.26775

3. Simons, D. G. Clinical and Etiological Update of Myofascial Pain from Trigger Points. Journal of Musculoskeletal Pain 4:93–122, 1996. 10.1300/J094v04n01_07

4. Do, T. P., G. F. Heldarskard, L. T. Kolding, J. Hvedstrup, and H. W. Schytz. Myofascial trigger points in migraine and tension-type headache. J Headache Pain 19:84, 2018. 10.1186/s10194-018-0913-8

5. Simons, D. G., J. G. Travell, and L. S. Simons. Travell & Simons’ myofascial pain and dysfunction: the trigger point manual. Baltimore: Williams & Wilkins, 1999, 1 pp.

6. Pawlukiewicz, M., M. Kochan, P. Niewiadomy, K. Szuścik-Niewiadomy, J. Taradaj, P. Król, and M. T. Kuszewski. Fascial Manipulation Method Is Effective in the Treatment of Myofascial Pain, but the Treatment Protocol Matters: A Randomised Control Trial—Preliminary Report. Journal of Clinical Medicine 11:4546, 2022. 10.3390/jcm11154546

7. van den Dolder, P. A., P. H. Ferreira, and K. M. Refshauge. Effectiveness of Soft Tissue Massage for Nonspecific Shoulder Pain: Randomized Controlled Trial. Phys Ther 95:1467–1477, 2015. 10.2522/ptj.20140350

8. Bingölbali, Ö., C. Taşkaya, H. Alkan, and Ö. Altındağ. The effectiveness of deep tissue massage on pain, trigger point, disability, range of motion and quality of life in individuals with myofascial pain syndrome. Somatosens Mot Res 41:11–17, 2024. 10.1080/08990220.2023.2165054

9. Raja G, P., S. Bhat, R. Gangavelli, A. Prabhu, A. Stecco, C. Pirri, V. Jaganathan, and C. Fernández-de-Las-Peñas. Effectiveness of Deep Cervical Fascial Manipulation® and Sequential Yoga Poses on Pain and Function in Individuals with Mechanical Neck Pain: A Randomised Controlled Trial. Life 13:2173, 2023. 10.3390/life13112173

10. Isaji, Y., D. Sasaki, K. Okuyama, Y. Kurasawa, K. Suzuki, Y. Kon, and T. Kitagawa. Therapeutic mechanisms of fascia manipulation: A scoping review. J Back Musculoskelet Rehabil 38:1267–1276, 2025. 10.1177/10538127251341828

11. Kodama, Y., S. Masuda, T. Ohmori, A. Kanamaru, M. Tanaka, T. Sakaguchi, and M. Nakagawa. Response to Mechanical Properties and Physiological Challenges of Fascia: Diagnosis and Rehabilitative Therapeutic Intervention for Myofascial System Disorders. Bioengineering (Basel) 10:474, 2023. 10.3390/bioengineering10040474

12. American Physical Therapy Associationat <https://www.apta.org/>

13. Keter, D. L., J. A. Bent, J. E. Bialosky, C. A. Courtney, J. E. Esteves, M. Funabashi, S. J. Howarth, H. S. Injeyan, A. M. Mazzieri, C. Glissmann Nim, and C. E. Cook. An international consensus on gaps in mechanisms of forced-based manipulation research: findings from a nominal group technique. J Man Manip Ther 32:111–117, 2024. 10.1080/10669817.2023.2262336

14. Gulick, D. T. Instrument-assisted soft tissue mobilization increases myofascial trigger point pain threshold. J Bodyw Mov Ther 22:341–345, 2018. 10.1016/j.jbmt.2017.10.012

15. Cumplido-Trasmonte, C., P. Fernández-González, I. M. Alguacil-Diego, and F. Molina-Rueda. Manual therapy in adults with tension-type headache: A systematic review. Neurologia (Engl Ed) 36:537–547, 2021. 10.1016/j.nrleng.2017.12.005

16. Jull, G., P. Trott, H. Potter, G. Zito, K. Niere, D. Shirley, J. Emberson, I. Marschner, and C. Richardson. A Randomized Controlled Trial of Exercise and Manipulative Therapy for Cervicogenic Headache. Spine 27:1835, 2002.

17. Adams, R., B. White, and C. Beckett. The Effects of Massage Therapy on Pain Management in the Acute Care Setting. Int J Ther Massage Bodywork 3:4–11, 2010.

18. Karels, C. H., W. Polling, S. M. A. Bierma-Zeinstra, A. Burdorf, A. P. Verhagen, and B. W. Koes. Treatment of arm, neck, and/or shoulder complaints in physical therapy practice. Spine (Phila Pa 1976) 31:E584–589, 2006. 10.1097/01.brs.0000229229.54704.77

19. Cheatham, S. W., R. Baker, and E. Kreiswirth. Instrument Assisted Soft Tissue Mobilization: A commentary on clinical practice guidelines for rehabilitation professionals. Int J Sports Phys Ther 14:670–682, 2019.

20. Berger, S. E., and A. T. Baria. Assessing Pain Research: A Narrative Review of Emerging Pain Methods, Their Technosocial Implications, and Opportunities for Multidisciplinary Approaches. Front Pain Res (Lausanne) 3:896276, 2022. 10.3389/fpain.2022.896276

21. Engell, S., J. J. Triano, J. R. Fox, H. M. Langevin, and E. E. Konofagou. Differential displacement of soft tissue layers from manual therapy loading. Clinical Biomechanics 33:66–72, 2016. 10.1016/j.clinbiomech.2016.02.011

22. Vappou, J., C. Maleke, and E. E. Konofagou. Quantitative viscoelastic parameters measured by harmonic motion imaging. Phys Med Biol 54:3579–3594, 2009. 10.1088/0031-9155/54/11/020

23. Chuang, L., C. Wu, and K. Lin. Reliability, Validity, and Responsiveness of Myotonometric Measurement of Muscle Tone, Elasticity, and Stiffness in Patients With Stroke. Archives of Physical Medicine and Rehabilitation 93:532–540, 2012. 10.1016/j.apmr.2011.09.014

24. Leonard, C. T., W. P. Deshner, J. W. Romo, E. S. Suoja, S. C. Fehrer, and E. L. Mikhailenok. Myotonometer Intra- and Interrater Reliabilities1. Archives of Physical Medicine and Rehabilitation 84:928–932, 2003. 10.1016/S0003-9993(03)00006-6

25. Romano, A., D. Staber, A. Grimm, C. Kronlage, and J. Marquetand. Limitations of Muscle Ultrasound Shear Wave Elastography for Clinical Routine-Positioning and Muscle Selection. Sensors (Basel) 21:8490, 2021. 10.3390/s21248490

26. Stecco, C., C. Fede, V. Macchi, A. Porzionato, L. Petrelli, C. Biz, R. Stern, and R. De Caro. The fasciacytes: A new cell devoted to fascial gliding regulation. Clin Anat 31:667–676, 2018. 10.1002/ca.23072

27. Stecco, A., L. Bonaldi, C. G. Fontanella, C. Stecco, and C. Pirri. The Effect of Mechanical Stress on Hyaluronan Fragments’ Inflammatory Cascade: Clinical Implications. Life (Basel) 13:2277, 2023. 10.3390/life13122277

28. Nešporová, K., J. Matonohová, J. Husby, E. Toropitsyn, L. D. Stupecká, A. Husby, T. Suchánková Kleplová, A. Streďanská, M. Šimek, D. Nečas, M. Vrbka, R. Schleip, and V. Velebný. Injecting hyaluronan in the thoracolumbar fascia: A model study. Int J Biol Macromol 253:126879, 2023. 10.1016/j.ijbiomac.2023.126879

29. Bishop, J. H., J. R. Fox, R. Maple, C. Loretan, G. J. Badger, S. M. Henry, M. A. Vizzard, and H. M. Langevin. Ultrasound Evaluation of the Combined Effects of Thoracolumbar Fascia Injury and Movement Restriction in a Porcine Model. PLoS One 11:e0147393, 2016. 10.1371/journal.pone.0147393

30. Wilke, J., F. Krause, L. Vogt, and W. Banzer. What Is Evidence-Based About Myofascial Chains: A Systematic Review. Arch Phys Med Rehabil 97:454–461, 2016. 10.1016/j.apmr.2015.07.023

31. Kumbhare, D. A., A. H. Elzibak, and M. D. Noseworthy. Assessment of Myofascial Trigger Points Using Ultrasound. Am J Phys Med Rehabil 95:72–80, 2016. 10.1097/PHM.0000000000000376

32. Jones, J. M., W. Foster, C. R. Twomey, J. Burdge, O. M. Ahmed, T. D. Pereira, J. A. Wojick, G. Corder, J. B. Plotkin, and I. Abdus-Saboor. A machine-vision approach for automated pain measurement at millisecond timescales. eLife 9:e57258, 2020. 10.7554/eLife.57258

33. Hauser, S. C., S. McIntyre, A. Israr, H. Olausson, and G. J. Gerling. Uncovering Human-to-Human Physical Interactions that Underlie Emotional and Affective Touch Communication. , 2019. 10.1109/WHC.2019.8816169

34. Hauser, S. C., S. S. Nagi, S. McIntyre, A. Israr, H. Olausson, and G. J. Gerling. From Human-to-Human Touch to Peripheral Nerve Responses. , 2019. 10.1109/WHC.2019.8816113

35. Lo, C., S. Ting Chu, T. B. Penney, and A. Schirmer. 3D Hand-Motion Tracking and Bottom-Up Classification Sheds Light on the Physical Properties of Gentle Stroking. Neuroscience , 2020. 10.1016/j.neuroscience.2020.09.037

36. Oikonomidis, I., N. Kyriazis, and A. Argyros. Efficient model-based 3D tracking of hand articulations using Kinect., 2011. 10.5244/C.25.101

37. Sridhar, S., F. Mueller, A. Oulasvirta, and C. Theobalt. Fast and robust hand tracking using detection-guided optimization. , 2015. 10.1109/CVPR.2015.7298941

38. Vonstad, E. K., X. Su, B. Vereijken, K. Bach, and J. H. Nilsen. Comparison of a Deep Learning-Based Pose Estimation System to Marker-Based and Kinect Systems in Exergaming for Balance Training. Sensors 20:6940, 2020. 10.3390/s20236940

39. Zhou, Y., M. Habermann, W. Xu, I. Habibie, C. Theobalt, and F. Xu. Monocular Real-Time Hand Shape and Motion Capture Using Multi-Modal Data. , 2020.at <https://openaccess.thecvf.com/content_CVPR_2020/html/Zhou_Monocular_Real-Time_Hand_Shape_and_Motion_Capture_Using_Multi-Modal_Data_CVPR_2020_paper.html>

40. Yan, Y., J. M. Goodman, D. D. Moore, S. A. Solla, and S. J. Bensmaia. Unexpected complexity of everyday manual behaviors. Nat Commun 11:3564, 2020. 10.1038/s41467-020-17404-0

41. Arteaga, M. V., J. C. Castiblanco, I. F. Mondragon, J. D. Colorado, and C. Alvarado-Rojas. EMG-driven hand model based on the classification of individual finger movements. Biomedical Signal Processing and Control 58:101834, 2020. 10.1016/j.bspc.2019.101834

42. Maycock, J., T. Rohlig, M. Schroder, M. Botsch, and H. Ritter. Fully automatic optical motion tracking using an inverse kinematics approach. , 2015. 10.1109/HUMANOIDS.2015.7363590

43. Delhaye, B. P., E. Jarocka, A. Barrea, J.-L. Thonnard, B. Edin, and P. Lefèvre. High-resolution imaging of skin deformation shows that afferents from human fingertips signal slip onset. eLife 10:e64679, 2021. 10.7554/eLife.64679

44. Delhaye, B., A. Barrea, B. B. Edin, P. Lefèvre, and J.-L. Thonnard. Surface strain measurements of fingertip skin under shearing. Journal of The Royal Society Interface 13:20150874, 2016. 10.1098/rsif.2015.0874

45. Dzidek, B., S. Bochereau, S. A. Johnson, V. Hayward, and M. J. Adams. Why pens have rubbery grips. PNAS 114:10864–10869, 2017. 10.1073/pnas.1706233114

46. Li, B., S. C. Hauser, and G. J. Gerling. Faster Indentation Influences Skin Deformation To Reduce Tactile Discriminability of Compliant Objects. IEEE Transactions on Haptics 16:215–227, 2023. 10.1109/TOH.2023.3253256

47. Hauser, S. C., and G. J. Gerling. Imaging the 3-D Deformation of the Finger Pad When Interacting with Compliant Materials. IEEE Haptics Symp 2018:7–13, 2018. 10.1109/HAPTICS.2018.8357145

48. Hu, X., R. Maiti, X. Liu, L. C. Gerhardt, Z. S. Lee, R. Byers, S. E. Franklin, R. Lewis, S. J. Matcher, and M. J. Carré. Skin surface and sub-surface strain and deformation imaging using optical coherence tomography and digital image correlation. , 2016. 10.1117/12.2212361

49. Maiti, R., L.-C. Gerhardt, Z. S. Lee, R. A. Byers, D. Woods, J. A. Sanz-Herrera, S. E. Franklin, R. Lewis, S. J. Matcher, and M. J. Carré. In vivo measurement of skin surface strain and sub-surface layer deformation induced by natural tissue stretching. J Mech Behav Biomed Mater 62:556–569, 2016. 10.1016/j.jmbbm.2016.05.035

50. Liu, X., R. Maiti, Z. H. Lu, M. J. Carré, S. J. Matcher, and R. Lewis. New Non-invasive Techniques to Quantify Skin Surface Strain and Sub-surface Layer Deformation of Finger-pad during Sliding. Biotribology 12:52–58, 2017. 10.1016/j.biotri.2017.07.001

51. Lee, Z. S., R. Maiti, M. J. Carré, and R. Lewis. Morphology of a human finger pad during sliding against a grooved plate: A pilot study. Biotribology 21:100114, 2020. 10.1016/j.biotri.2019.100114

52. Xu, Z., J. D. Cruz, C. Fthenakis, and C. Saliou. A novel method to measure skin mechanical properties with three-dimensional digital image correlation. Skin Research and Technology 25:60–67, 2019. 10.1111/srt.12596

53. Kao, A. R., T. M. Loghmani, and G. J. Gerling. Strain-based biomarkers at the skin surface differentiate asymmetries in soft tissue mobility associated with myofascial pain. Journal of the Mechanical Behavior of Biomedical Materials 172:107175, 2025. 10.1016/j.jmbbm.2025.107175

54. Morgan, D. Principles of Soft Tissue Treatment. Journal of Manual & Manipulative Therapy 2:63–65, 1994. 10.1179/jmt.1994.2.2.63

55. Langevin, H. M. Fascia Mobility, Proprioception, and Myofascial Pain. Life (Basel) 11:668, 2021. 10.3390/life11070668

56. Rezaei, M., S. S. Nagi, C. Xu, S. McIntyre, H. Olausson, and G. J. Gerling. Thin Films on the Skin, but not Frictional Agents, Attenuate the Percept of Pleasantness to Brushed Stimuli. , 2021. 10.1109/WHC49131.2021.9517259

57. AMTAat <https://www.amtamassage.org/>

58. Bhattacharjee, A., S. Y. P. Chien, S. Anwar, and M. T. Loghmani. Quantifiable Soft Tissue Manipulation (QSTM^TM^) - A novel modality to improve clinical manual therapy with objective metrics. Annu Int Conf IEEE Eng Med Biol Soc 2021:4961–4964, 2021. 10.1109/EMBC46164.2021.9629616

59. Bhattacharjee, A., S. Anwar, S. Chien, and M. T. Loghmani. A Handheld Quantifiable Soft Tissue Manipulation Device for Tracking Real-Time Dispersive Force-Motion Patterns to Characterize Manual Therapy Treatment. IEEE Trans Biomed Eng PP:, 2022. 10.1109/TBME.2022.3222124

60. Burke, J., D. J. Buchberger, M. T. Carey-Loghmani, P. E. Dougherty, D. S. Greco, and J. D. Dishman. A Pilot Study Comparing Two Manual Therapy Interventions for Carpal Tunnel Syndrome. Journal of Manipulative and Physiological Therapeutics 30:50–61, 2007. 10.1016/j.jmpt.2006.11.014

61. Carey-Loghmani, M. Graston Technique Training Manual: Module 1. 1998.

62. Sutton, M., W. Wolters, W. Peters, W. Ranson, and S. McNeill. Determination of displacements using an improved digital correlation method. Image and Vision Computing 1:133–139, 1983. 10.1016/0262-8856(83)90064-1

63. Bruck, H. A., S. R. McNeill, M. A. Sutton, and W. H. Peters. Digital image correlation using Newton-Raphson method of partial differential correction. Experimental Mechanics 29:261–267, 1989. 10.1007/BF02321405

64. Solav, D., K. M. Moerman, A. M. Jaeger, K. Genovese, and H. M. Herr. MultiDIC: An Open-Source Toolbox for Multi-View 3D Digital Image Correlation. IEEE Access 6:30520–30535, 2018. 10.1109/ACCESS.2018.2843725

65. Blaber, J., B. Adair, and A. Antoniou. Ncorr: Open-Source 2D Digital Image Correlation Matlab Software. Exp Mech 55:1105–1122, 2015. 10.1007/s11340-015-0009-1

66. Pourahmadi, M., J. Dommerholt, C. Fernández-de-Las-Peñas, B. W. Koes, M. A. Mohseni-Bandpei, M. A. Mansournia, S. Delavari, A. Keshtkar, and M. Bahramian. Dry Needling for the Treatment of Tension-Type, Cervicogenic, or Migraine Headaches: A Systematic Review and Meta-Analysis. Physical Therapy 101:pzab068, 2021. 10.1093/ptj/pzab068

67. Kelley, M. J., M. A. Shaffer, J. E. Kuhn, L. A. Michener, A. L. Seitz, T. L. Uhl, J. J. Godges, and P. McClure. Shoulder Pain and Mobility Deficits: Adhesive Capsulitis. Journal of Orthopaedic & Sports Physical Therapy 43:A1–A31, 2013. 10.2519/jospt.2013.0302

68. DeVocht, J. W., J. G. Pickar, and D. G. Wilder. Spinal Manipulation Alters Electromyographic Activity of Paraspinal Muscles: A Descriptive Study. Journal of Manipulative and Physiological Therapeutics 28:465–471, 2005. 10.1016/j.jmpt.2005.07.002

69. Clopper, C. J., and E. S. Pearson. THE USE OF CONFIDENCE OR FIDUCIAL LIMITS ILLUSTRATED IN THE CASE OF THE BINOMIAL. Biometrika 26:404–413, 1934. 10.1093/biomet/26.4.404

70. Bernal-Utrera, C., J. J. Gonzalez-Gerez, E. Anarte-Lazo, and C. Rodriguez-Blanco. Manual therapy versus therapeutic exercise in non-specific chronic neck pain: a randomized controlled trial. Trials 21:682, 2020. 10.1186/s13063-020-04610-w

71. Nadal-Nicolás, Y., J. Á. Rubio-Arias, M. Martínez-Olcina, C. Reche-García, M. Hernández-García, and A. Martínez-Rodríguez. Effects of Manual Therapy on Fatigue, Pain, and Psychological Aspects in Women with Fibromyalgia. Int J Environ Res Public Health 17:4611, 2020. 10.3390/ijerph17124611

72. Hvedstrup, J., L. T. Kolding, M. Ashina, and H. W. Schytz. Increased neck muscle stiffness in migraine patients with ictal neck pain: A shear wave elastography study. Cephalalgia 40:565–574, 2020. 10.1177/0333102420919998

73. Loghmani M, T., and S. Bane. Instrument-assisted Soft Tissue Manipulation: Evidence for its Emerging Efficacy. J Nov Physiother s3:, 2016. 10.4172/2165-7025.S3-012

74. Abu Taleb, W., A. Rehan Youssef, and A. Saleh. The effectiveness of manual versus algometer pressure release techniques for treating active myofascial trigger points of the upper trapezius. J Bodyw Mov Ther 20:863–869, 2016. 10.1016/j.jbmt.2016.02.008

75. Kablan, N., N. Alaca, and Y. Tatar. Comparison of the Immediate Effect of Petrissage Massage and Manual Lymph Drainage Following Exercise on Biomechanical and Viscoelastic Properties of the Rectus Femoris Muscle in Women. J Sport Rehabil 30:725–730, 2021. 10.1123/jsr.2020-0276

76. Mahmood, T., A. Abrar, M. M. Atif, W. Mahmood, and F. Batool. Effect of instrument-assisted soft tissue mobilization versus myofascial release therapy for pain, mobility, and disability in chronic low backache patients: a quasi-experimental study. Anaesthesia, Pain & Intensive Care 28:452–458, 2024. 10.35975/apic.v28i3.2459

77. Anderst, W. J., T. Gale, C. LeVasseur, S. Raj, K. Gongaware, and M. Schneider. Intervertebral kinematics of the cervical spine before, during, and after high-velocity low-amplitude manipulation. Spine J 18:2333–2342, 2018. 10.1016/j.spinee.2018.07.026

78. Langevin, H. M., D. Stevens-Tuttle, J. R. Fox, G. J. Badger, N. A. Bouffard, M. H. Krag, J. Wu, and S. M. Henry. Ultrasound evidence of altered lumbar connective tissue structure in human subjects with chronic low back pain. BMC Musculoskeletal Disorders 10:151, 2009. 10.1186/1471-2474-10-151

79. Langevin, H. M., J. R. Fox, C. Koptiuch, G. J. Badger, A. C. Greenan-Naumann, N. A. Bouffard, E. E. Konofagou, W.-N. Lee, J. J. Triano, and S. M. Henry. Reduced thoracolumbar fascia shear strain in human chronic low back pain. BMC Musculoskelet Disord 12:203, 2011. 10.1186/1471-2474-12-203

80. Tomita, N., M.-H. Roy-Cardinal, B. Chayer, S. Daher, A. Attiya, A. Boulanger, N. Gaudreault, G. Cloutier, and N. J. Bureau. Thoracolumbar fascia ultrasound shear strain differs between low back pain and asymptomatic individuals: expanding the evidence. Insights Imaging 16:18, 2025. 10.1186/s13244-024-01895-2

81. Speicher, T. E., N. M. Selkow, and A. J. Warren. Manual Therapy Improves Immediate Blood Flow and Tissue Fiber Alignment of the Forearm Extensors. J Phys Med Rehabil Volume 4:28–36, 2022. 10.33696/rehabilitation.4.029

82. Jiménez-del-Barrio, S., A. Cadellans-Arróniz, L. Ceballos-Laita, E. Estébanez-de-Miguel, C. López-de-Celis, E. Bueno-Gracia, and A. Pérez-Bellmunt. The effectiveness of manual therapy on pain, physical function, and nerve conduction studies in carpal tunnel syndrome patients: a systematic review and meta-analysis. International Orthopaedics (SICOT) 46:301–312, 2022. 10.1007/s00264-021-05272-2

83. Davidson, C. J., L. R. Ganion, G. M. Gehlsen, B. Verhoestra, J. E. Roepke, and T. L. Sevier. Rat tendon morphologic and functional changes resulting from soft tissue mobilization. Med Sci Sports Exerc 29:313–319, 1997. 10.1097/00005768-199703000-00005

84. Stecco, A., M. Gesi, C. Stecco, and R. Stern. Fascial Components of the Myofascial Pain Syndrome. Curr Pain Headache Rep 17:352, 2013. 10.1007/s11916-013-0352-9

85. Fede, C., A. Angelini, R. Stern, V. Macchi, A. Porzionato, P. Ruggieri, R. De Caro, and C. Stecco. Quantification of hyaluronan in human fasciae: variations with function and anatomical site. Journal of Anatomy 233:552–556, 2018. 10.1111/joa.12866

86. Courbot, O., and A. Elosegui-Artola. The role of extracellular matrix viscoelasticity in development and disease. NPJ Biol Phys Mech 2:10, 2025. 10.1038/s44341-025-00014-6

87. Ikeda, N., S. Otsuka, Y. Kawanishi, and Y. Kawakami. Effects of Instrument-assisted Soft Tissue Mobilization on Musculoskeletal Properties. Med Sci Sports Exerc 51:2166–2172, 2019. 10.1249/MSS.0000000000002035

88. Zügel, M., C. N. Maganaris, J. Wilke, K. Jurkat-Rott, W. Klingler, S. C. Wearing, T. Findley, M. F. Barbe, J. M. Steinacker, A. Vleeming, W. Bloch, R. Schleip, and P. W. Hodges. Fascial tissue research in sports medicine: from molecules to tissue adaptation, injury and diagnostics: consensus statement. Br J Sports Med 52:1497, 2018. 10.1136/bjsports-2018-099308

89. Findley, T., H. Chaudhry, A. Stecco, and M. Roman. Fascia research – A narrative review. Journal of Bodywork and Movement Therapies 16:67–75, 2012. 10.1016/j.jbmt.2011.09.004

90. Loghmani, M. T., and S. J. Warden. Instrument-Assisted Cross-Fiber Massage Accelerates Knee Ligament Healing. Journal of Orthopaedic & Sports Physical Therapy 39:506–514, 2009. 10.2519/jospt.2009.2997

91. Iwatsuki, H., Y. Ikuta, and K. Shinoda. Deep Friction Massage on the Masticatory Muscles in Stroke Patients Increases Biting Force. Journal of Physical Therapy Science 13:17–20, 2001. 10.1589/jpts.13.17

92. Coronado, R. A., C. W. Gay, J. E. Bialosky, G. D. Carnaby, M. D. Bishop, and S. Z. George. Changes in pain sensitivity following spinal manipulation: a systematic review and meta-analysis. J Electromyogr Kinesiol 22:752–767, 2012. 10.1016/j.jelekin.2011.12.013

93. McCormack, J. R., F. B. Underwood, E. J. Slaven, and T. A. Cappaert. Eccentric Exercise Versus Eccentric Exercise and Soft Tissue Treatment (Astym) in the Management of Insertional Achilles Tendinopathy: A Randomized Controlled Trial. Sports Health 8:230–237, 2016. 10.1177/1941738116631498

94. Papa, J. A. Two cases of work-related lateral epicondylopathy treated with Graston Technique® and conservative rehabilitation. J Can Chiropr Assoc 56:192–200, 2012.

95. Fernández-Pérez, A. M., C. Villaverde-Gutiérrez, A. Mora-Sánchez, C. Alonso-Blanco, M. Sterling, and C. Fernández-de-las-Peñas. Muscle Trigger Points, Pressure Pain Threshold, and Cervical Range of Motion in Patients With High Level of Disability Related to Acute Whiplash Injury. Journal of Orthopaedic & Sports Physical Therapy 42:634–641, 2012. 10.2519/jospt.2012.4117

96. Andersen, H., L. Arendt-Nielsen, B. Danneskiold-SamsØe, and T. Graven-Nielsen. Pressure pain sensitivity and hardness along human normal and sensitized muscle. Somatosensory & Motor Research 23:97–109, 2006. 10.1080/08990220600856255

97. Ransone, J. W., J. Schmidt, S. K. Crawford, and J. Walker. Effect of manual compressive therapy on latent myofascial trigger point pressure pain thresholds. Journal of Bodywork and Movement Therapies 23:792–798, 2019. 10.1016/j.jbmt.2019.06.011

98. Dommerholt, J., C. Bron, and J. Franssen. Myofascial Trigger Points: An Evidence-Informed Review. Journal of Manual & Manipulative Therapy 14:203–221, 2006. 10.1179/106698106790819991

99. Schleip, R., and D. G. Müller. Training principles for fascial connective tissues: scientific foundation and suggested practical applications. J Bodyw Mov Ther 17:103–115, 2013. 10.1016/j.jbmt.2012.06.007

100. Pratt, R. L. Hyaluronan and the Fascial Frontier. Int J Mol Sci 22:6845, 2021. 10.3390/ijms22136845

101. Ganjaei, K. G., J. W. Ray, B. Waite, and K. J. Burnham. The Fascial System in Musculoskeletal Function and Myofascial Pain. Curr Phys Med Rehabil Rep 8:364–372, 2020. 10.1007/s40141-020-00302-3

102. Reeves, J. L., B. Jaeger, and S. B. Graff-Radford. Reliability of the pressure algometer as a measure of myofascial trigger point sensitivity. Pain 24:313–321, 1986. 10.1016/0304-3959(86)90117-X

103. Coulter, I. D., C. Crawford, E. L. Hurwitz, H. Vernon, R. Khorsan, M. Suttorp Booth, and P. M. Herman. Manipulation and mobilization for treating chronic low back pain: a systematic review and meta-analysis. Spine J 18:866–879, 2018. 10.1016/j.spinee.2018.01.013

104. Coppieters, M. W., K. H. Stappaerts, L. L. Wouters, and K. Janssens. The Immediate Effects of a Cervical Lateral Glide Treatment Technique in Patients With Neurogenic Cervicobrachial Pain. Journal of Orthopaedic & Sports Physical Therapy 33:369–378, 2003. 10.2519/jospt.2003.33.7.369

105. Anwer, S., A. Alghadir, H. Zafar, and J.-M. Brismée. Effects of orthopaedic manual therapy in knee osteoarthritis: a systematic review and meta-analysis. Physiotherapy 104:264–276, 2018. 10.1016/j.physio.2018.05.003

106. Fernández-de-las-Peñas, C., J. Cleland, M. Palacios-Ceña, S. Fuensalida-Novo, J. A. Pareja, and C. Alonso-Blanco. The Effectiveness of Manual Therapy Versus Surgery on Self-reported Function, Cervical Range of Motion, and Pinch Grip Force in Carpal Tunnel Syndrome: A Randomized Clinical Trial. Journal of Orthopaedic & Sports Physical Therapy 47:151–161, 2017. 10.2519/jospt.2017.7090

107. Bini, P., D. Hohenschurz-Schmidt, V. Masullo, D. Pitt, and J. Draper-Rodi. The effectiveness of manual and exercise therapy on headache intensity and frequency among patients with cervicogenic headache: a systematic review and meta-analysis. Chiropractic & Manual Therapies 30:49, 2022. 10.1186/s12998-022-00459-9

108. Wu, Z., Y. Zhu, W. Xu, J. Liang, Y. Guan, and X. Xu. Analysis of Biomechanical Properties of the Lumbar Extensor Myofascia in Elderly Patients with Chronic Low Back Pain and That in Healthy People. BioMed Research International 2020:7649157, 2020. 10.1155/2020/7649157

109. Lee, Y., M. Kim, and H. Lee. The Measurement of Stiffness for Major Muscles with Shear Wave Elastography and Myoton: A Quantitative Analysis Study. Diagnostics (Basel) 11:524, 2021. 10.3390/diagnostics11030524

110. Vuorenmaa, A. S., E. M. K. Siitama, and K. S. Mäkelä. Inter-operator and inter-device reproducibility of shear wave elastography in healthy muscle tissues. J Appl Clin Med Phys 23:e13717, 2022. 10.1002/acm2.13717

111. Valera-Calero, J. A., S. Sánchez-Jorge, J. Buffet-García, U. Varol, G. M. Gallego-Sendarrubias, and J. Álvarez-González. Is Shear-Wave Elastography a Clinical Severity Indicator of Myofascial Pain Syndrome? An Observational Study. Journal of Clinical Medicine 10:2895, 2021. 10.3390/jcm10132895

112. Eriksson Crommert, M., L. Lacourpaille, L. J. Heales, K. Tucker, and F. Hug. Massage induces an immediate, albeit short-term, reduction in muscle stiffness. Scandinavian Journal of Medicine & Science in Sports 25:e490–e496, 2015. 10.1111/sms.12341

113. McNulty, W. H., R. N. Gevirtz, D. R. Hubbard, and G. M. Berkoff. Needle electromyographic evaluation of trigger point response to a psychological stressor. Psychophysiology 31:313–316, 1994. 10.1111/j.1469-8986.1994.tb02220.x

114. Sikdar, S., J. Srbely, J. Shah, Y. Assefa, A. Stecco, S. DeStefano, M. Imamura, and L. H. Gerber. A model for personalized diagnostics for non-specific low back pain: the role of the myofascial unit. Front Pain Res (Lausanne) 4:1237802, 2023. 10.3389/fpain.2023.1237802

115. Glassman, G. E., L. Dellalana, E. R. Tkaczyk, I. M. Esteve, J. Y. Huang, A. Cronin, J. R. Patrinely, S. Etemad, P. E. Assi, S. Ridner, A. J. Forte, S. A. Kassis, and G. Perdikis. Measuring Biomechanical Properties Using a Noninvasive Myoton Device for Lymphedema Detection and Tracking: A Pilot Study. Eplasty 22:e54, 2022.

116. Gerwin, R. D., S. Shannon, C. Z. Hong, D. Hubbard, and R. Gevirtz. Interrater reliability in myofascial trigger point examination. Pain 69:65–73, 1997. 10.1016/s0304-3959(96)03248-4

117. Rathbone, A. T. L., L. Grosman-Rimon, and D. A. Kumbhare. Interrater Agreement of Manual Palpation for Identification of Myofascial Trigger Points: A Systematic Review and Meta-Analysis. Clin J Pain 33:715–729, 2017. 10.1097/AJP.0000000000000459

