## Supplemental Figures for "Detecting intervention-specific change in soft tissue mobility aligned with regional pain through optically measured skin surface strains"

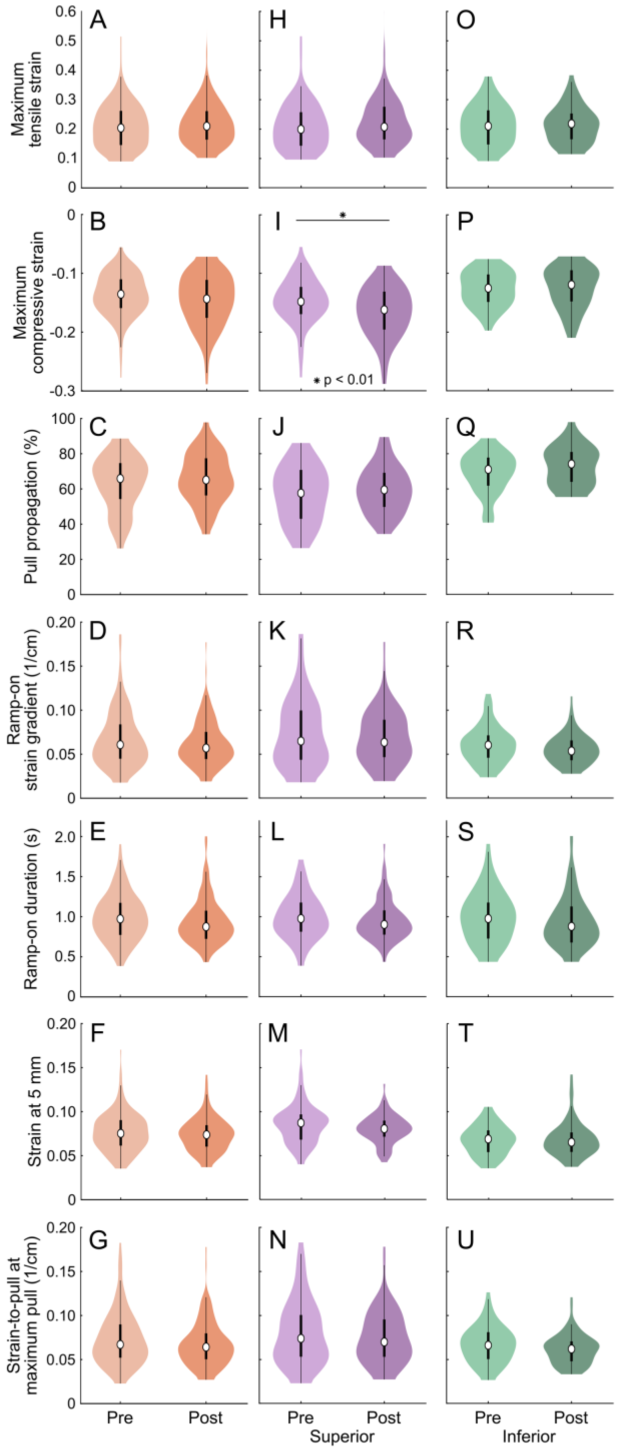


**Supplemental Fig. S1** Participant population trends: pre- to post-intervention mobility changes, presented both in aggregate and separated by pull direction (superior, inferior). (a)-(g) Biomarker distribution across all participants with sides and directions aggregated, separated by intervention condition, and analyzed using linear mixed-effects (LME) models (α = 0.01). Post-intervention, four biomarkers were significantly changed (zero shown). (h)-(n) Aggregate data isolated for the superior pull direction, separated by intervention condition, and analyzed using LME models (α = 0.01). Significant changes were observed in four biomarkers (one shown: (i)). Specifically, (i) maximum compressive strain decreased post-intervention ($t(170) = -3.10$, $p < 0.01$) indicating more compression. (o)-(u) Aggregate data isolated for the inferior pull direction, separated by intervention condition, and analyzed using LME models (α = 0.01). Significant changes were observed in one biomarker, ramp-on pull velocity (not shown)

**Supplemental Table S1** Participant population trends: pre- to post-intervention mobility change statistics, presented both in aggregate and separated by pull direction

| | Unpaired | | Paired | | | |
|---|---|---|---|---|---|---|
| Biomarker | *p* | *t* | *p* | *t* | df | Change |
| Aggregate | | | | | | |
| Maximum tensile strain | > 0.01 | 0.99 | > 0.01 | 1.30 | 342 | 5.9E-03 |
| Maximum pull | **< 0.01** | 3.18 | **< 0.01** | 4.64 | 342 | 3.3 |
| Maximum compressive strain | > 0.01 | -2.11 | **< 0.01** | -2.64 | 342 | -8.0E-03 |
| Maximum far region displacement | **< 0.01** | 3.03 | **< 0.01** | 4.44 | 342 | 2.7 |
| Pull propagation | > 0.01 | 2.00 | **< 0.01** | 2.63 | 341 | -0.8 |
| Ramp-on pull velocity | **< 0.01** | 4.29 | **< 0.01** | 5.33 | 341 | 8.4 |
| Ramp-on strain gradient | > 0.01 | -1.72 | > 0.01 | -2.58 | 341 | -3.8E-04 |
| Ramp-on duration | > 0.01 | -1.72 | > 0.01 | -1.89 | 341 | -0.1 |
| Strain at 5 mm | > 0.01 | -1.26 | > 0.01 | -1.43 | 342 | -1.5E-03 |
| Strain-to-pull at maximum pull | > 0.01 | -2.00 | **< 0.01** | -3.06 | 342 | -2.9E-04 |
| Total gross deformation | **< 0.01** | 3.35 | **< 0.01** | 4.18 | 342 | 0.6 |
| Superior pull direction | | | | | | |
| Maximum tensile strain | > 0.01 | 1.17 | > 0.01 | 2.23 | 170 | 8.0E-03 |
| Maximum pull | **< 0.01** | 2.72 | **< 0.01** | 7.20 | 170 | 4.9 |
| Maximum compressive strain | **< 0.01** | -3.10 | **< 0.01** | -5.52 | 170 | -1.4E-02 |
| Maximum far region displacement | > 0.01 | 2.22 | **< 0.01** | 6.01 | 170 | 2.3 |
| Pull propagation | > 0.01 | 1.15 | > 0.01 | 2.23 | 169 | 1.9 |
| Ramp-on pull velocity | **< 0.01** | 3.15 | **< 0.01** | 5.53 | 169 | 8.1 |
| Ramp-on strain gradient | > 0.01 | -1.10 | > 0.01 | -1.99 | 169 | -1.4E-04 |
| Ramp-on duration | > 0.01 | -1.12 | > 0.01 | -1.12 | 169 | -0.1 |
| Strain at 5 mm | > 0.01 | -2.04 | > 0.01 | -2.55 | 170 | -6.7E-03 |
| Strain-to-pull at maximum pull | > 0.01 | -1.24 | > 0.01 | -2.28 | 170 | -4.0E-04 |
| Total gross deformation | **< 0.01** | 3.04 | **< 0.01** | 5.73 | 170 | 0.6 |
| Inferior pull direction | | | | | | |
| Maximum tensile strain | > 0.01 | 0.11 | > 0.01 | 0.16 | 170 | 7.9E-03 |
| Maximum pull | > 0.01 | 1.92 | **< 0.01** | 3.87 | 170 | 2.0 |
| Maximum compressive strain | > 0.01 | 0.16 | > 0.01 | 0.21 | 170 | 5.8E-03 |
| Maximum far region displacement | > 0.01 | 2.35 | **< 0.01** | 5.63 | 170 | 2.1 |
| Pull propagation | > 0.01 | 2.21 | **< 0.01** | 5.17 | 170 | 3.1 |
| Ramp-on pull velocity | **< 0.01** | 3.06 | **< 0.01** | 3.96 | 170 | 10.3 |
| Ramp-on strain gradient | > 0.01 | -1.70 | **< 0.01** | -3.13 | 170 | -6.6E-04 |
| Ramp-on duration | > 0.01 | -1.31 | > 0.01 | -1.57 | 170 | -0.1 |
| Strain at 5 mm | > 0.01 | 0.18 | > 0.01 | 0.22 | 170 | -3.7E-03 |
| Strain-to-pull at maximum pull | > 0.01 | -1.99 | **< 0.01** | -3.97 | 170 | -4.2E-04 |
| Total gross deformation | > 0.01 | 1.80 | **< 0.01** | 2.76 | 170 | 0.5 |

Linear mixed-effects model results ($\alpha = 0.01$) are reported for all pulls combined (aggregate – top) and separated by pull direction (superior – middle; inferior – bottom). The table lists *t* and *p* values, degrees of freedom (*df*), and median pre-to-post changes for both participant-unpaired and participant-paired analyses. Unpaired analyses identified four significant biomarkers (bolded, green) in aggregate (maximum pull, maximum far region displacement, ramp-on pull velocity, and total gross deformation), four in the superior direction, and one in the inferior direction. Paired analyses retained these effects while revealing additional significant biomarkers, totaling seven biomarkers in aggregate, five in the superior direction, and seven in the inferior direction. The greater number of significant findings in the paired analyses reflects increased sensitivity after accounting for individual baseline differences.

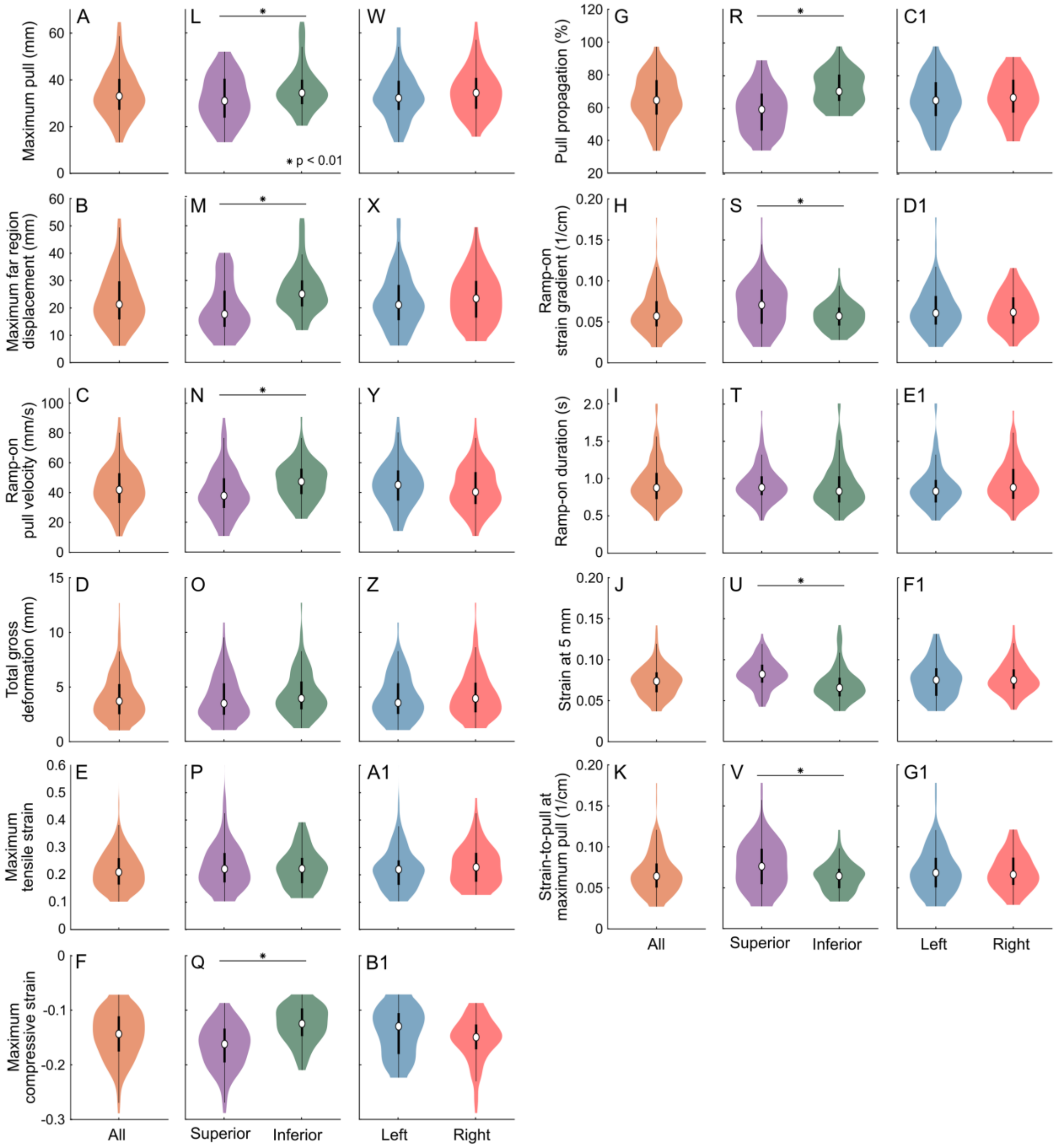


**Supplemental Fig. S2** Participant population trends: post-intervention asymmetries in tissue mobility given pull in superior vs. inferior direction and assessment of left vs. right body side. (a)-(k) Biomarker distribution across all participants with sides and directions aggregated, analyzed using linear mixed-effects (LME) models (α = 0.01). In (a) the tissue allowed for 33.1 ± 10.2 mm (mean ± SD) of maximum pull which in (b) produced 21.4 ± 9.7 mm of maximum far region displacement. Similarly, in (c), the ramp-on pull velocity of the tissue measured 42.1 ± 14.9 mm/s post-intervention. (l)-(v) Aggregate data post-intervention separated by pull direction and analyzed using LME models (α = 0.01). Significant directional asymmetries were observed in the same eight biomarkers as pre-intervention (l-n, q-s, u, v). (w)-(g1) Aggregate data post-intervention separated by body side and analyzed using LME models (α = 0.01). No significant bilateral asymmetries were found for any of the biomarkers

**Supplemental Table S2** Individual trends: pre-intervention asymmetries in tissue mobility given assessment of left vs. right body side

| Mobility asymmetries between **left** and **right body sides** in **pre-intervention** assessment (right – left; %) | | | | | | | | | | | | | | | | | | | |
|---|---|---|---|---|---|---|---|---|---|---|---|---|---|---|---|---|---|---|---|
| Participant | 1 | 2 | 3 | 4 | 5 | 6 | 7 | 8 | 9 | 10 | 11 | 12 | 13 | 14 | 15 | 16 | 17 | 18 | 19 |
| Superior pull direction | | | | | | | | | | | | | | | | | | | |
| Maximum pull | -6.0 | **-28.7** | -3.9 | -7.5 | 17.7 | -14.8 | 1.2 | **34.5** | 12.8 | 24.8 | -13.1 | 8.8 | **31.5** | 14.7 | 4.9 | -2.3 | -14.2 | 4.5 | -5.1 |
| Far region displacement | 16.1 | -4.7 | **30.2** | **47.5** | 17.1 | -0.2 | 15.6 | **61.4** | 19.2 | **40.7** | **-26.0** | -2.4 | **67.2** | 19.6 | -7.8 | -16.6 | -13.8 | 11.3 | -7.2 |
| Ramp-on pull velocity | -15.5 | **-28.9** | **-32.6** | **35.3** | -6.3 | **-50.2** | -17.5 | -12.8 | 3.8 | 23.4 | -5.4 | 19.0 | 10.2 | 12.9 | 22.3 | -23.4 | -7.9 | -6.0 | -23.0 |
| Total gross deformation | -10.9 | **-60.0** | -2.5 | 24.1 | **29.2** | -22.8 | -6.0 | **29.3** | 14.3 | **42.3** | -13.5 | **44.0** | -0.1 | 21.4 | **36.2** | 15.3 | -14.0 | 9.9 | 18.7 |
| Inferior pull direction | | | | | | | | | | | | | | | | | | | |
| Maximum pull | **29.3** | -22.7 | 8.4 | **35.5** | 12.8 | 1.6 | -0.5 | 2.0 | 8.6 | 3.4 | 7.9 | -12.3 | **42.6** | **32.3** | 17.1 | **-31.1** | -4.3 | 2.5 | 6.2 |
| Far region displacement | **28.8** | **-38.9** | -14.5 | **34.5** | 6.6 | 7.6 | 0.3 | 7.5 | 6.7 | 16.8 | 0.0 | -7.5 | **31.0** | **32.2** | 16.4 | **-30.2** | -0.7 | 6.5 | -1.7 |
| Ramp-on pull velocity | **26.4** | -19.2 | 9.1 | **49.2** | -7.4 | **-27.2** | -24.4 | -3.2 | -15.8 | -18.5 | 22.3 | 3.0 | **36.0** | **30.4** | 11.4 | **-44.2** | 20.7 | 2.2 | 5.7 |
| Total gross deformation | **38.4** | **-37.8** | **51.8** | **65.2** | 21.1 | 10.0 | -6.8 | -13.2 | 5.5 | -3.9 | 24.1 | -23.0 | **58.0** | **48.4** | **38.1** | **-49.5** | -12.2 | 1.7 | 15.5 |
| Pre-intervention total | **4** | **-5** | 1 | **6** | 1 | **-2** | 0 | **3** | 0 | **2** | -1 | 1 | **6** | **4** | **2** | **-4** | 0 | 0 | 0 |
| More mobile side | R | L | | R | | L | | R | | R | | | R | R | R | L | | | |
| Alignment between DIC mobility and self-reported pain (**M**—Match; **NM**—No match) | | | | | | | | | | | | | | | | | | | |
| Less mobile side | L | R | | L | | R | | L | | L | | | L | L | L | R | | | |
| More painful side | L | R | L | L | | | | L | | | | R | L | L | L | R | | | |
| 80% agreement | M | M | NM | M | | | | M | | | | NM | M | M | M | M | | | |

Pre-intervention mobility asymmetries were identified using the four selected biomarkers applied across both pull directions. Biomarker differences were computed as right minus left, such that positive values indicate greater mobility on the right body side and negative values indicate greater mobility on the left body side (*Methods—Statistical Analysis*). The table reports, for each participant and pull direction (superior—top block; inferior—middle block), the number of selected biomarker comparisons exceeding the predefined threshold, with counts summed across pull directions to yield eight total comparisons per participant (four biomarkers x two directions). Comparisons exceeding threshold are bolded, with blue shading indicating positive values and red shading indicating negative values. A participant was classified as more mobile on the right side ("R") if they had a sum of two or more comparisons exceeding threshold (blue), or more mobile on the left side ("L") if they had a sum of negative two or more comparisons exceeding threshold (red). DIC identified ten participants (52.6%) as having bilaterally asymmetrical mobility pre-intervention: seven were more mobile on their right (P1, P4, P8, P10, P13, P14, P15) and three were more mobile on their left (P2, P6, P16). In eight of these cases (80%; P1, P2, P4, P8, P13, P14, P15, P16), the body side measured as less mobile by DIC corresponded with the more painful side by self-report, indicated by "M" for match or "NM" for no match

**Supplemental Table S3** Sensitivity of individual-level mobility classification to threshold choice (20%, 25%, 30%)

Participant changes between **pre-intervention** and **post-intervention** at different **cutoff thresholds** (%)

| Participant | 1 | 2 | 3 | 4 | 5 | 6 | 7 | 8 | 9 | 10 | 11 | 12 | 13 | 14 | 15 | 16 | 17 | 18 | 19 | **Treated participant classifications** (n=17) | | |
|---|---|---|---|---|---|---|---|---|---|---|---|---|---|---|---|---|---|---|---|---|---|---|
| Threshold | Left body side | | | | | | | | | | | | | | | | | | | Improved | Declined | No change |
| 20% | **3** | **2** | **3** | **8** | 1 | **2** | **2** | **4** | **-7** | 1 | **2** | **3** | **5** | **7** | **7** | -1 | **5** | **3** | 1 | 16 (94.1%) | 1 (5.9%) | 0 (0%) |
| 25% | **3** | **2** | **3** | **7** | 0 | 1 | 0 | **4** | **-5** | 1 | **2** | **2** | **5** | **5** | **5** | 0 | **3** | **3** | 0 | 15 (88.2%) | 1 (5.9%) | 1 (5.9%) |
| 30% | **3** | **3** | **2** | **7** | 0 | 1 | 0 | **4** | **-2** | 1 | 1 | **2** | **4** | **3** | **4** | 0 | **2** | **3** | 0 | 13 (76.5%) | 1 (5.9%) | 3 (17.6%) |
| Threshold | Right body side | | | | | | | | | | | | | | | | | | | | | |
| 20% | **3** | **3** | **-2** | **2** | 1 | **4** | **4** | **2** | **-3** | 1 | **4** | **4** | **3** | **4** | -1 | **4** | **6** | **3** | **2** | Net counts (positive − negative) of biomarker comparisons exceeding threshold (8 possible per body side: 4 biomarkers × 2 pull directions). Shaded cells indicate a body side is improved (≥2; blue) or declined (≤−2; red). | | |
| 25% | **3** | **3** | -1 | **2** | 0 | **4** | **2** | 1 | -1 | 1 | **3** | **3** | **2** | 1 | -1 | **3** | **4** | 1 | 1 | | | |
| 30% | **3** | **3** | -1 | **2** | 0 | **2** | 0 | 1 | -1 | 1 | **2** | **3** | 1 | 1 | 0 | 1 | **4** | 0 | 0 | | | |

A sensitivity analysis for post-intervention mobility changes was conducted across three classification thresholds. The table reports, for each participant and body side, the number of biomarker comparisons exceeding each of three thresholds: 20%, 25% (predefined), and 30%. Values are net counts (positive − negative) of biomarker comparisons exceeding the given threshold, out of 8 possible per body side (4 biomarkers × 2 pull directions), separated by body side (left—top section; right—bottom section). Counts exceeding threshold (≥2 or ≤−2) are bolded, with blue shading indicating a body side classified as improved and red shading indicating a body side classified as declined. The 25% rows match the published manuscript Table 1. Results were consistent with the original findings: among the 17 treated participants, the proportion classified as improved ranged from 76.5% (30% threshold) to 94.1% (20% threshold), compared to 88.2% at our reported 25% threshold, with the direction and general magnitude of the effect preserved throughout. The single participant showing declined mobility (P9) was consistent across all three thresholds. Of the 15 participants originally classified as improved at 25%, 12 remained so regardless of threshold; 2 participants (P7, P16) shifted to "no change" at the more conservative threshold (30%) tested, while 1 participant (P19) shifted from "no change" to "improved" at the less conservative threshold (20%) tested. Both untreated participants (P5, P10) remained classified as showing no mobility change across all three thresholds.

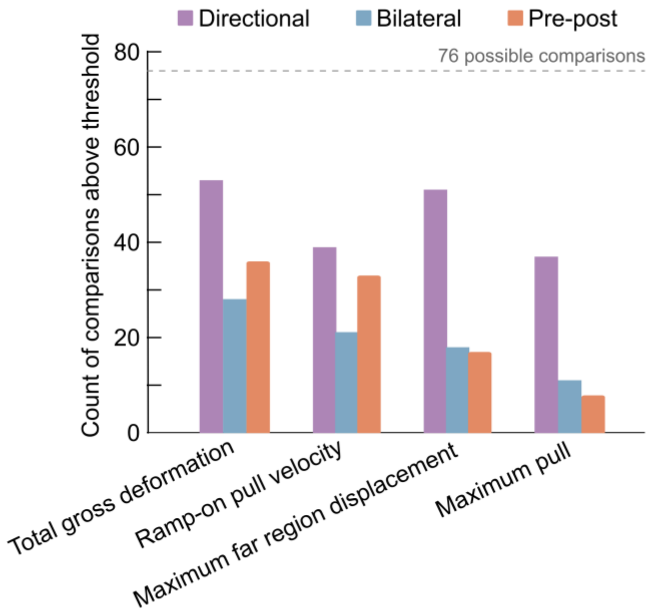


**Supplemental Fig. S3** Biomarker sensitivity. Across all participants (n=19), bars indicate the number of threshold-exceeding comparisons for each of the four selected biomarkers, separated by category (directional—purple, bilateral—blue, pre-post—orange). With four comparisons per category, each biomarker had 76 opportunities for detection, or 228 opportunities total. Of the 352 total comparisons exceeding threshold, the highest frequency was in the directional category (51.1%), followed by pre-post (26.7%) and bilateral (22.2%). Total gross deformation and ramp-on pull velocity showed the greatest sensitivity, with 117 (51.3%) and 93 (40.8%) total detections, respectively. Maximum far region displacement showed moderate sensitivity (86 comparisons; 37.7%), while maximum pull yielded the fewest detections (56 comparisons; 24.6%)
